# Targeted long-read sequencing enables comprehensive analysis of the genetic and epigenetic landscape of inherited myopathies

**DOI:** 10.64898/2025.12.06.25340828

**Authors:** Dennis Yeow, Andre LM Reis, Igor Stevanovski, Neysa Njo, Laura I Rudaks, Bianca R Grosz, Joanne S Sy, Leah Kemp, Sanjog R Chintalaphani, Michael Chin, Marion Stoll, Danqing Zhu, Christina Liang, Katrina Morris, Andrew Hannaford, Ehsan Shandiz, Kate E Ahmad, Shadi El-Wahsh, Stephen W Reddel, Robert Boland-Freitas, Roula Ghaoui, Stephanie Barnes, Jonathan Sturm, Anna Willard, Mahi Jasinarachchi, Simon Hawke, Neil G Simon, Lisa Worgan, David Manser, Michel Tchan, Neil C Griffith, Ryan L Davis, Michael C Fahey, Carolyn M Sue, Pamela A McCombe, Karl Ng, Marina L Kennerson, Pak Leng Cheong, Kishore R Kumar, Ira W Deveson

## Abstract

Inherited myopathies are a group of disorders with diverse and complex genetic aetiologies. The causative genetic variants vary widely in type, size and sequence context, encompassing small sequence variants and large structural variants in protein-coding genes, mitochondrial variants, repeat expansions, and more complex events, such as the chromosome 4 D4Z4 macrosatellite contraction and hypomethylation that causes facioscapulohumeral muscular dystrophy (FSHD). This poses a challenge for analysis with next generation sequencing and other molecular methods. This is further compounded by phenotypic variability between patients and phenotypic overlap of different forms of genetic myopathy. To accelerate myopathy research and improve diagnosis we have developed a targeted long-read sequencing assay and an integrated bioinformatics analysis framework that captures the full suite of genes, variants and epigenetic signatures currently implicated in all forms of inherited myopathy. Applying this to a cohort of 53 myopathy patients with and without previous genetic diagnoses, we demonstrate the analytical validity of our approach, as well as its improved accuracy and resolution compared to existing methods – especially for FSHD. Our LRS assay identified an array of new information about the genetic and epigenetic landscape of inherited myopathies and provided new diagnoses for 29% of patients who had remained undiagnosed following clinical genetic testing. In our cohort, FSHD and oculopharyngodistal myopathies were the most common new diagnoses that were missed or mis-diagnosed by standard clinical genetic testing. Our new method constitutes a single streamlined assay for comprehensive genetic and epigenetic characterisation of inherited myopathies.

## Introduction

Genetic myopathies (inherited muscle diseases) are a large group of heterogeneous disorders that affect an estimated 1 in 1700–4500 people.^1,2^ Molecular characterisation of patients with genetic myopathies is essential to understanding the genetic basis of disease, developing new therapies, and providing patients with an accurate diagnosis, which facilitates disease-specific treatment, prognosis, genetic counselling, family planning, and access to clinical trials.^3^ However, the process is complicated by several inter-related challenges. The genetic myopathies exhibit high phenotypic heterogeneity, i.e., patients with the same genetic cause may present with different clinical phenotypes, disease severity and age of onset. There is also significant phenotypic overlap between different types of genetic myopathy, i.e., patients with different genetic causes may present with similar clinical features. Finally, the known genetic causes of myopathy are numerous and highly diverse, occurring in a large number of genes (>300 described to date) across the nuclear and mitochondrial genomes. These include single and multi-nucleotide variants (SNVs, MNVs), small insertions and deletions (indels), structural and copy number variants (SVs, CNVs), expanded short (STRs) and variable number tandem repeats (VNTRs), and contractions of the D4Z4 macrosatellite that may cause facioscapulohumeral muscular dystrophy (FSHD).^4^ Epigenetic regulation of gene promoters, STRs or the D4Z4 macrosatellite is also central to the disease mechanism in certain myopathies – methylation profiling can be informative for diagnosis and prognosis in these disorders.^4,5^

In the absence of a single assay that can capture all potential molecular causes, these genetic and clinical complexities limit our ability to diagnose genetic myopathies and understand their pathophysiology.^4^ While short read next-generation sequencing (NGS) provides high-throughput assessment of hundreds of myopathy genes, it has limited capacity to resolve repetitive or non-unique genetic loci, phase distantly located genetic variants, identify SVs/CNVs and other complex structural rearrangements, characterise repeat expansions/contractions, and detect epigenetic changes. To date, application of NGS to undiagnosed myopathy cohorts have resulted in diagnostic yields of 21–65%.^6–9^ Alternative, lower throughput, disorder-specific testing methodologies are required to diagnose myopathies refractory to NGS.^4,10^ For example, the common genetic myopathies myotonic dystrophy type 1 (DM1) and 2 (DM2) are caused by STR expansions in the *DMPK* and *CNBP* genes, and typically require loci-specific repeat-primed PCR (RP-PCR) or Southern blot (SB) for diagnosis.^11^ However, the rapidly increasing number of newly described genetic myopathies caused by STR expansions (e.g. oculopharyngodistal myopathies [OPDM]) lack readily accessible testing, as each newly described loci requires design and validation of a gene-specific RP-PCR or SB assay.^4^

FSHD, the third most common genetic myopathy, is especially complex. Genetic diagnosis of FSHD may involve a combination of SB to identify chromosome 4 D4Z4 macrosatellite repeat contractions and 4q haplotyping (A or B) that contribute to FSHD type 1 (FSHD1), NGS to identify SNVs or indels in genes that cause FSHD type 2 (FSHD2), and/or bisulphite sequencing (BSS) to detect D4Z4 hypomethylation, seen in both FSHD1 and FSHD2.^12^ Although SB was the gold standard for FSHD1 diagnosis, it is low throughput, labour intensive, imprecise and may generate false-positive and false-negative results as a result of uncommon, structurally complex D4Z4 alleles.^12^ Optical genome mapping and molecular combing are newer techniques that are superior to SB for detection of D4Z4 contractions and SVs, but neither identifies the sequence variants that cause FSHD2 or the abnormal D4Z4 methylation profile seen in FSHD.^4,13–15^ As a result, there is a great need for an integrated approach that fully resolves the genetic and epigenetic architecture of FSHD (and genetic myopathies in general) in order to better understand the molecular mechanisms of the disorder and provide patients with accurate, timely diagnoses.

Given these challenges, we reasoned that long-read sequencing (LRS), a newer sequencing technology that addresses many of the limitations of NGS, would be particularly well suited to the molecular characterisation of genetic myopathies.^4^ LRS enables detection of large, repetitive and/or complex genetic variants and mapping of reads across homologous and/or repetitive regions of the genome. It also readily phases distant variants to identify compound heterozygous events in the absence of parental sequencing data. Both of the currently available LRS platforms, Oxford Nanopore Technologies (ONT) and Pacific Biosciences (PacBio), are able to concurrently assess epigenetic changes without the need for additional sample processing.^16^ Therefore, by capturing all known genetic and epigenetic alterations in a single test, LRS may better navigate genotype/phenotype heterogeneity and improve genetic myopathy diagnosis.

We previously demonstrated the use of targeted LRS to simultaneously characterise all STR expansions implicated in neuromuscular disease.^17^ Here, we build on this approach by developing a targeted LRS assay and integrated bioinformatics framework to assess all known genetic/epigenetic causes of inherited myopathies. This approach delivers new genetic insights and diagnoses within a cohort of patients with suspected genetic myopathies who have previously received inconclusive or negative results via conventional clinical genetic testing. Our new method is the first example of a single comprehensive assay suitable for diagnostic investigation of all known forms of genetic myopathy.

## Results

### A targeted ONT LRS assay for genetic myopathies

We developed a targeted ONT LRS assay using adaptive sampling to enrich for a ∼60 Mbase target panel encompassing all loci implicated in genetic myopathies (*n* = 331 at the time of panel creation in March 2023). The panel included individual myopathy genes, myopathy-causing STR loci, the mitochondrial genome, and the subtelomeric regions of 4q and 10q relevant to FSHD (<u>Table S1</u>). We developed an efficient bioinformatics pipeline to detect, phase and annotate genetic variants of all types, and determine 5-methylcytosine (5mC) DNA methylation frequencies within genomic target regions (see **Methods**). We also developed a new bioinformatics software package, named **d4z4ling** (read: “dazzling”), that uses LRS data to resolve the genetic and epigenetic contributors to FSHD, described later.

We applied our LRS assay to 53 participants with genetic myopathies, including those with an established genetic diagnosis (*n* = 22; ‘Group 1’) (Fig 1A; <u>Tables 1</u> & <u>2</u>) and those with suspected genetic myopathies who remained genetically unsolved or incompletely solved after previous clinical genetic testing (*n* = 31; ‘Group 2’; <u>Fig. S1</u>). Participants in Group 2 had a mean disease duration (symptom onset to study recruitment) of 15.5 years. The cohort was processed at a rate of 1–3 participant samples per ONT flow cell, obtaining 18–250-fold average coverage and 16–48 kb read length N_50_ within genomic target regions (<u>Fig. S2A-C</u>). An average of 210–1,824-fold coverage was obtained for the mitochondrial genome (<u>Fig.</u> <u>S2D</u>).

**Figure 1.**
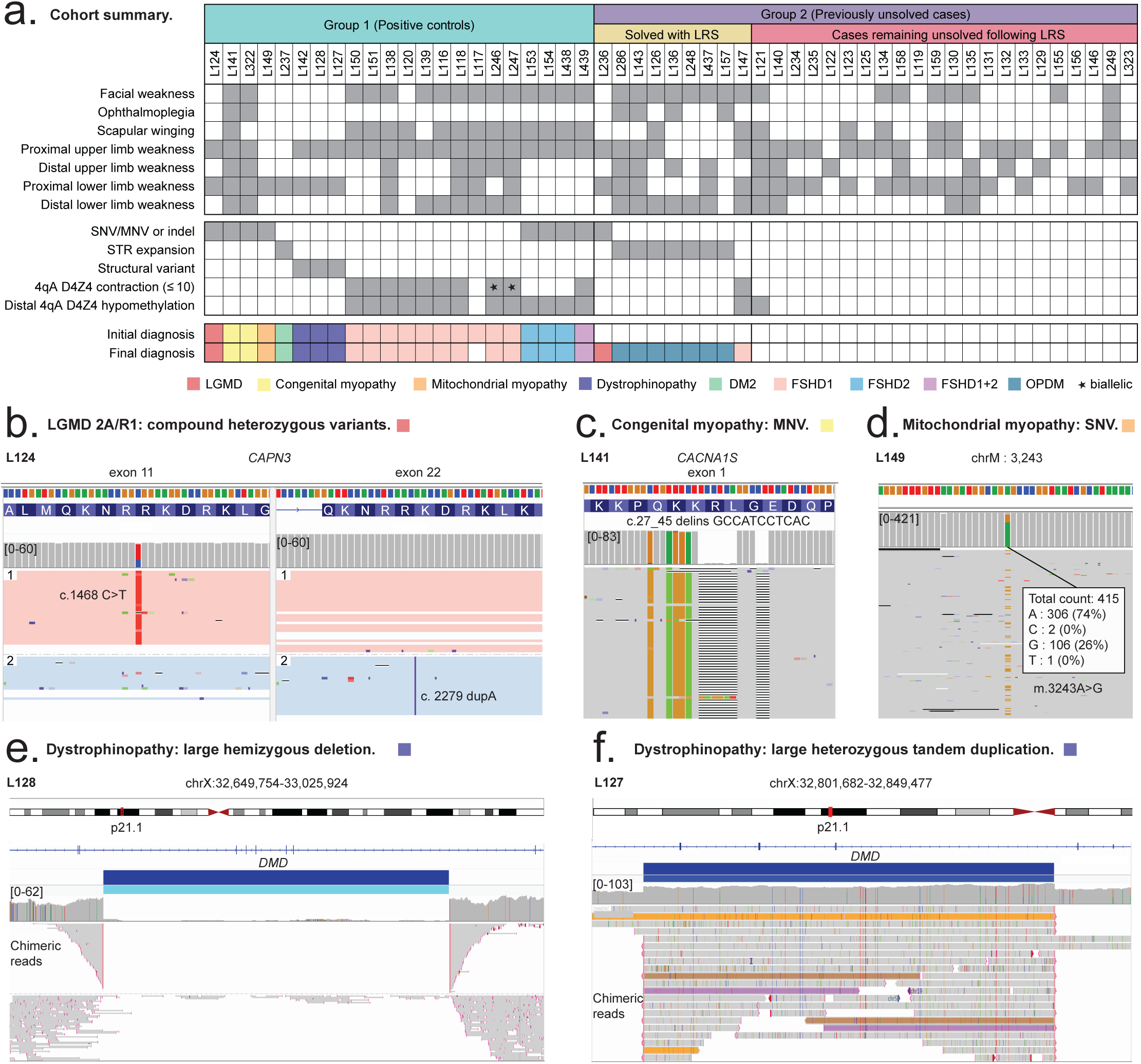
Cohort characteristics and examples of known genetic variants identified by ONT LRS. (**A**) Cohort summary with participants grouped into positive controls, newly solved cases following ONT LRS, and cases remaining unsolved following ONT LRS. The upper matrix summarises the distribution of muscle weakness and the lower matrix summarises the molecular findings contributing to each diagnosis. Grey boxes indicate the presence of a clinical feature or variant type. Coloured boxes in the final row denote diagnoses before and after ONT LRS. (**B**) Case L124, with a known diagnosis of LGMD 2A/R1, harbours a missense variant (c.1468C>T) in exon 11 and duplication (c.2279dupA) in exon 22 of *CAPN3*. ONT LRS was able to confirm the two variants were on different alleles (red & blue). (**C**) Case L141, with a known diagnosis of congenital myopathy 18, harbours a complex, homozygous, multinucleotide variant (c.27_45delinsGCCATCCTCAC) in exon 1 of *CACNA1S*. (**D)** Case L149, with a known diagnosis of m.3243A>G-related mitochondrial myopathy, harbours this variant with heteroplasmy of 26% in blood. (**E**) Case L128, with a known diagnosis of Duchenne muscular dystrophy, harbours a large deletion of *DMD* exons 3–7. ONT LRS generated chimeric reads spanning the deletion, allowing for identification of the deletion breakpoint and the presence of a small 9bp insertion at the deletion site: c.93+60377_650-16627delinsACTTGTTTG. (**F**) Case L127 is a manifesting carrier of a *DMD* exon 5–7 duplication. ONT LRS identified the deletion was in tandem (indicated by individual chimeric long reads [examples highlighted in purple and orange] that map across both sides of the duplicated interval) and flanked by a CT dinucleotide sequence: c.649+3234_649+3235ins[CT;264+2035_649+3234;CT].

Our LRS assay identified previously known causative variants in 21 of 22 participants in Group 1, including SNVs, MNVs, indels, mtDNA variants, STR expansions, SV/CNVs, and D4Z4 macrosatellite contractions, 4qA/B haplotypes and DNA hypomethylation (Fig. 1; <u>Tables 1</u> & <u>2</u>). The exception was case L117, for whom a previous diagnosis of FSHD1 was revoked based on findings from LRS data (see below). LRS also provided more complete genetic characterisation than available from previous testing in several cases. For example, the capacity to phase distantly located variants allowed for confirmation that pathogenic variants in *CAPN3* and *RYR1* were biallelic in two cases (L124, L322), in the absence of data from family members (Fig. 1B; <u>Fig. S3A</u>). In case L237, with a diagnosis of DM2, previous RP-PCR identified the presence of a pathogenic CCTG STR expansion in intron 1 of *CNBP* but was unable to size it accurately. LRS reads spanning the STR site confirmed an upper size of 2870 repeat copies, with some evidence of repeat length mosaicism (ranging from 2275–2870 repeats) (<u>Fig.S3B</u>). For three dystrophinopathy cases (L142, L128, L127) previously diagnosed using *DMD* multiplex ligation-dependent probe amplification (MLPA), LRS identified these CNVs and resolved precise CNV breakpoints and orientations (Fig. 1E-F; <u>Fig. S3C</u>). For example, in case L127, MLPA previously identified the presence of a duplication of *DMD* exons 5–7 but could not determine if the duplicated segment was in tandem, nor its orientation. LRS reads proved the duplication was in tandem and non-inverted (Fig. 1F). This is clinically relevant since extragenic (non-tandem) *DMD* duplications are not expected to cause a dystrophinopathy and are largely benign.^18^ Knowledge of exact breakpoints for large SVs/CNVs within *DMD* can also elucidate the mechanisms by which they arise (<u>Fig. S4</u>).^19^ For example, in case L128, both ends of the duplicated segment were found to be located within partially homologous LINE-1 retrotransposon elements within introns 4 and 7, indicating this duplication may have arisen from LINE-LINE-mediated nonallelic homologous recombination.^20^

In Group 2 our LRS assay led to a new genetic diagnosis in 29% (9/31) (Table 3), and revealed an array of new genetic and epigenetic findings, explored below. LRS also either identified new suspicious variants of uncertain significance (VUS) requiring further validation, or provided phasing data for previously identified suspicious VUS in several of the cases that remained genetically unsolved (<u>Supplementary Note1</u>). Overall, these results demonstrated the validity of our experimental and analytical approaches, and the capacity of LRS to improve genetic characterisation of patients with myopathies.

**Table 1.**
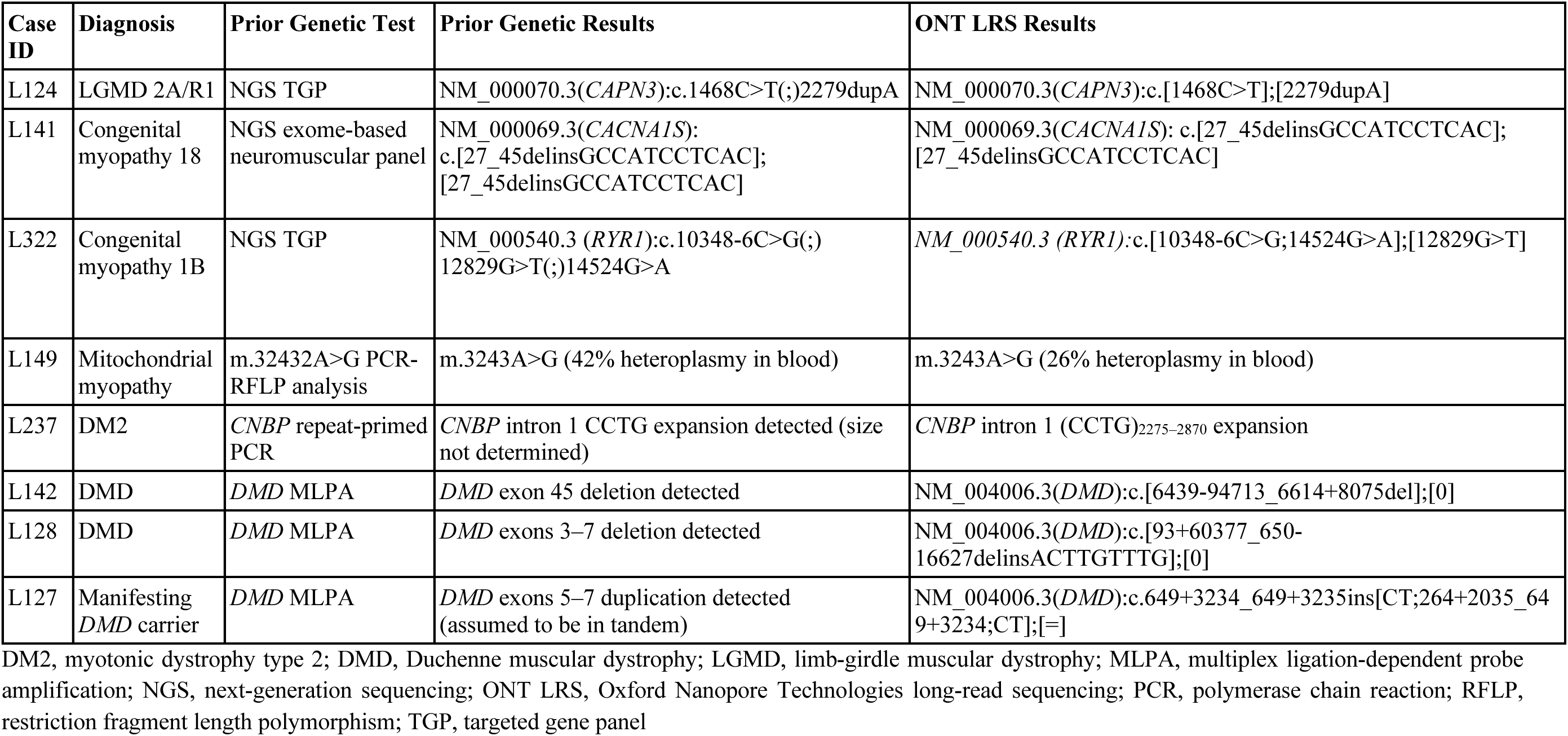
ONT LRS results in myopathy patients with prior known genetic diagnoses.

**Table 2.** Genetic & epigenetic variants from ONT LRS in a cohort of patients with prior diagnoses of FSHD.

| Case ID | Diagnosis Prior to ONT LRS | Prior Genetic Results |  | ONT LRS Results |  |  |  | Final Diagnosis Following ONT LRS |
| --- | --- | --- | --- | --- | --- | --- | --- | --- |
|  |  | Estimated D4Z4 Copy Number from SB* | Other Genetic Results | D4Z4 Copy Number & Contraction Haplotype | Methylation Pattern | Mean (S.D.) SSLP Length on Contracted Allele, bp | Other Genetic Result |  |
| L150 | FSHD1 | 5 | - | 5 on 4qA | FSHD1 | 160.83 (1.77) | - | FSHD1 |
| L151 | FSHD1 | 9 | - | 10 on 4qA | FSHD1 | 161.4 (1.53) | - | FSHD1 |
| L138 | FSHD1 | 4 | 4-copy D4Z4 array on 4qA allele using haplotype-specific SB | 4 on 4qA | FSHD1 | 160.6 (2.27) | - | FSHD1 |
| L120 | FSHD1 | 6 | - | 5 on 4qA | FSHD1 | 163.5 (2.52) | - | FSHD1 |
| L139 | FSHD1 | 4 | 4-copy D4Z4 array on 4qA allele using haplotype-specific SB | 4 on 4qA | FSHD1 | 161 (2.00) | - | FSHD1 |
| L116 | FSHD1 | 6 | - | 7 on 4qA | FSHD1 | 160.5 (2.17) | - | FSHD1 |
| L117 | FSHD1 | 10 | 10-copy D4Z4 array on 4qA allele using haplotype-specific SB | 10 on 10qA; no 4qA contraction | Normal | 174.91 (1.14) | - | None; FSHD diagnosis revoked |
| L118 | FSHD1 | 5 | - | 6 on 4qA | FSHD1 | 160.86 (2.12) | - | FSHD1 |
| L246<sup>†</sup> | FSHD1 | 7 & 9 | FSHD1 methylation pattern on BSS | 7 & 9 both on 4qA | FSHD1 (biallelic) | 161.24 (2.54), 160.8 (3.49) | - | FSHD1 |
| L247<sup>†</sup> | FSHD1 | 6 & 8 | FSHD2 methylation pattern on BSS | 7 & 9 both on 4qA | FSHD1 (biallelic) | 160.22 (2.32), 160.17 (2.62) | - | FSHD1 |
| L153<sup>‡</sup> | FSHD2 | >10 | NM_015295.3(*SMCHD1*):c.[4347-129A>G];[=] on srWGS; FSHD2 methylation pattern on BSS | In *cis* D4Z4 duplication on 4qA | FSHD2 | - | NM_015295.3(*SMCHD1*):c.[4347-129A>G];[=] | FSHD2 |
| L154<sup>‡</sup> | FSHD2 | >10 | NM_015295.3(*SMCHD1*):c.[4347-129A>G];[=] on Sanger sequencing; FSHD2 methylation pattern on BSS | In *cis* D4Z4 duplication on 4qA | FSHD2 | - | NM_015295.3(*SMCHD1*):c.[4347-129A>G];[=] | FSHD2 |
| L438<sup>‡</sup> | FSHD2 | >10 | Assumed to have same *SMCHD1* variant as daughters (L153, L154) | In *cis* D4Z4 duplication on 4qA | FSHD2 | - | NM_015295.3(*SMCHD1*):c.[4347-129A>G];[=] | FSHD2 |
|  |  | Estimated D4Z4 Copy Number from SB* | Other Genetic Results | D4Z4 Copy Number & Contraction Haplotype | Methylation Pattern | Mean (S.D.) SSLP Length on Contracted Allele, bp | Other Genetic Result |  |
| L150 | FSHD1 | 5 | - | 5 on 4qA | FSHD1 | 160.83 (1.77) | - | FSHD1 |
| L439 | FSHD1+2 | 7 | NM_015295.3(*SMCHD1*):c.2604-10_2615del on exome sequencing | 7 on 4qA | FSHD1+2 | 158.63 (4.32) | NM_015295.3(*SMCHD1*):c.[2604-10_2615del];[=] | FSHD1+2 |

BSS, bisulphite sequencing; FSHD, facioscapulohumeral muscular dystrophy; ONT LRS, Oxford Nanopore Technologies long-read sequencing; S.D., standard deviation; SB, Southern blot; srWGS, short-read whole genome sequencing; SSLP, short sequence length polymorphism
\*SB performed on *EcoRI/BlnI* double digested DNA separated by pulse-field gel electrophoresis hybridised with p13E-11 probe
†siblings
‡siblings (L153, L154) and mother (L438)

**Table 3.**
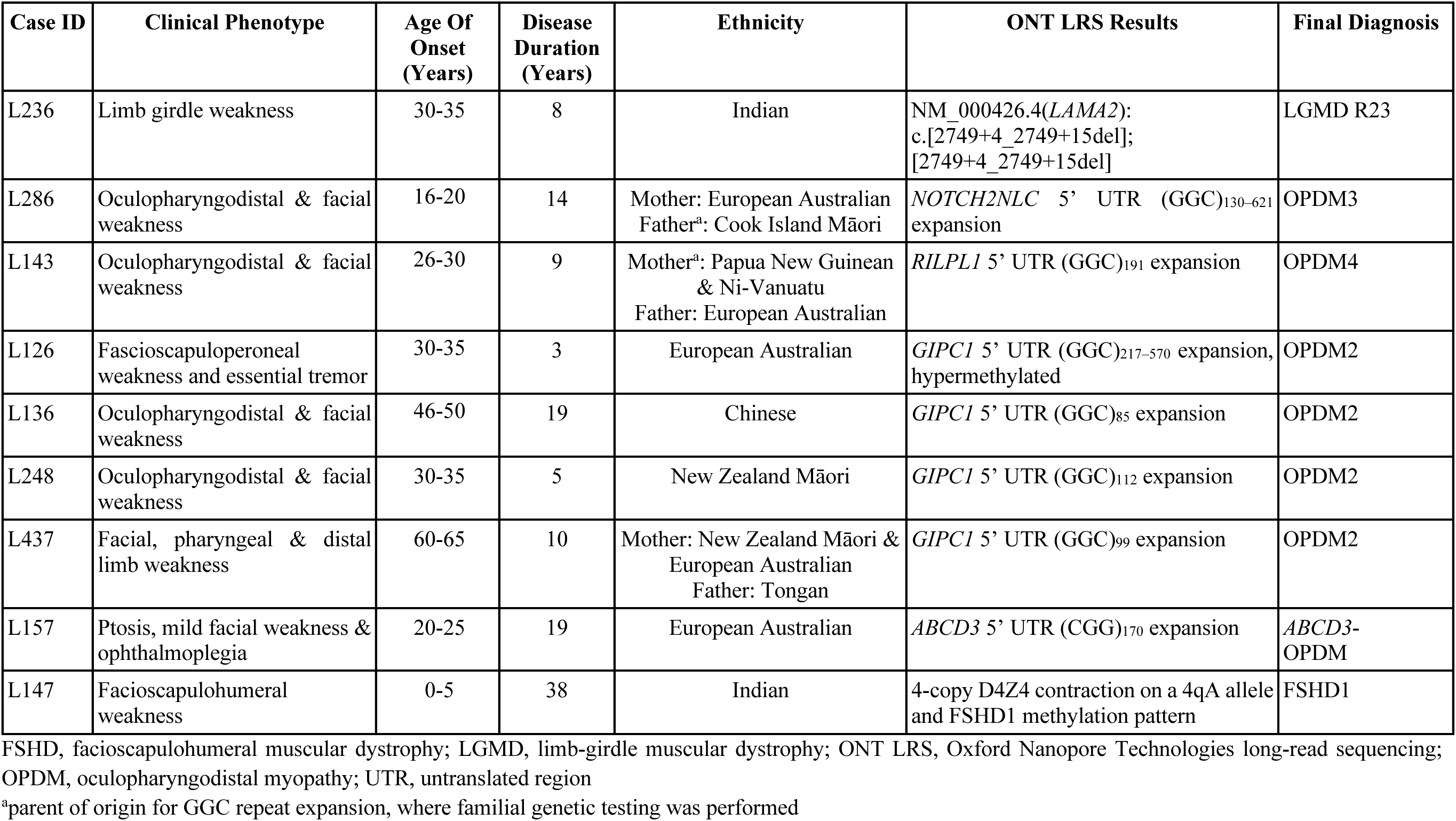
New myopathy diagnoses identified by ONT LRS.

### A new method for FSHD analysis

An integrated molecular and computational method for analysis of patients with FSHD forms a key component of our assay. Our targeted LRS panel captures the subtelomeric regions of chromosomes 4q, containing the D4Z4 macrosatellite that is contracted in FSHD1, and 10q, containing a highly homologous D4Z4 macrosatellite that does not contribute to FSHD. We developed a DNA shearing and size selection method to maximise yields of ONT reads long enough to span a contracted, pathogenic D4Z4 haplotype (10 copies; >33 kb; see **Methods**). Our new software package, **d4z4ling** (https://github.com/neysa-15/d4z4ling), analyses reads that fully and/or partially span the D4Z4 repeat array to determine D4Z4 copy number, DNA methylation status, haplotype (4qA, 4qB, 10qA, 10qB), and identify other supporting sequence features on each D4Z4 copy in a given patient (Fig. 2A<u>&B</u>). The genes involved in FSHD2 (*SMCHD1*, *DNMT3B*, *LRIF1*) are simultaneously captured and profiled for functionally relevant variants. Our LRS assay therefore captures all genomic and epigenomic features relevant to FSHD.

**Figure 2.**
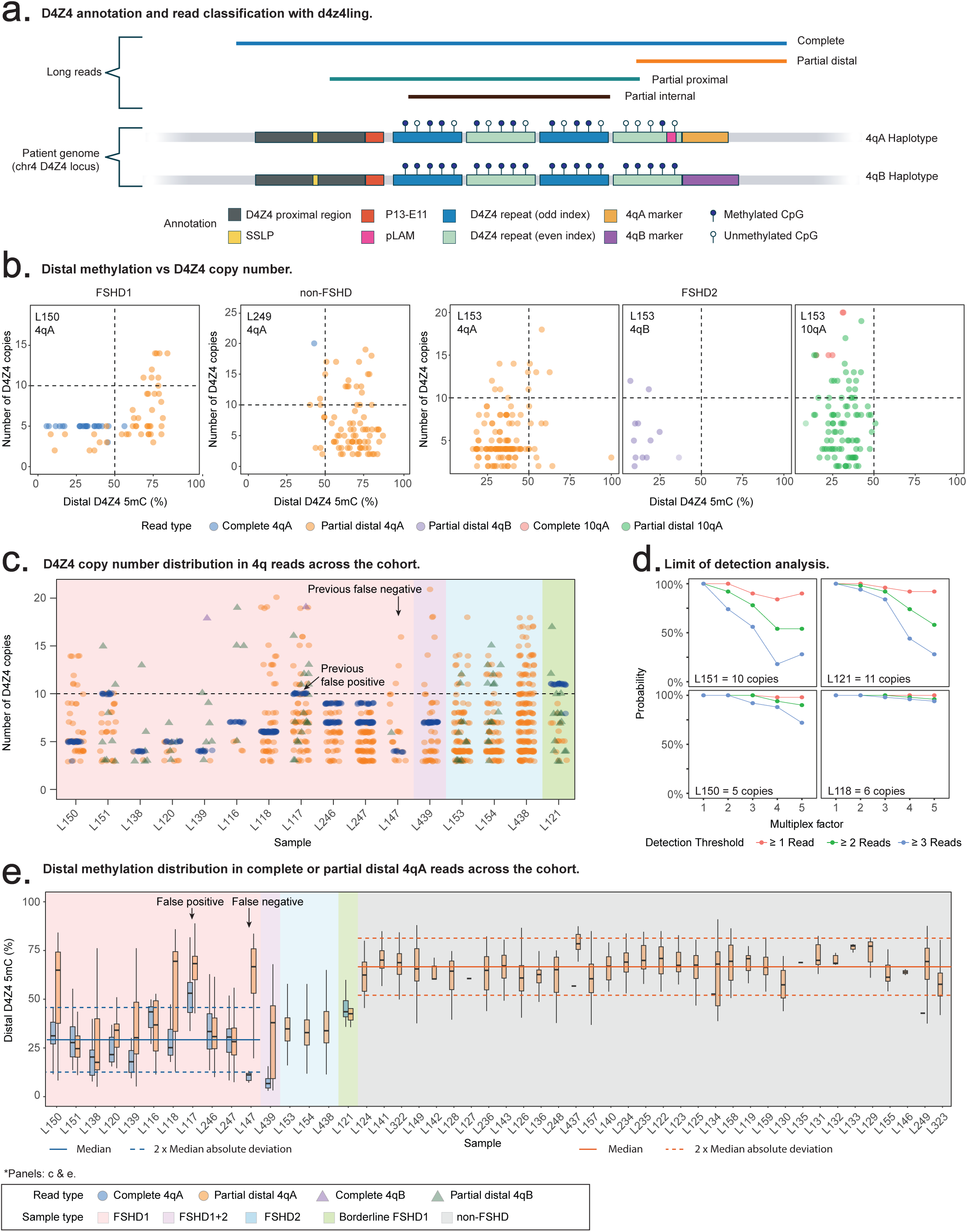
Targeted ONT LRS characterises the genetic and epigenetic architecture of FSHD. (**A**) Schematic of D4Z4 annotation and read classification at the chromosome 4q D4Z4 locus. Long reads are classified as complete, partial distal, partial proximal, or partial internal depending on their alignment span across the entire repeat array. Each read is annotated with key features, including proximal elements (SSLP, p13-E11), D4Z4 repeat units, the distal pLAM region, and 4qA/4qB haplotype-specific markers. CpG methylation is measured along the array for each read, with the fraction of methylation at the distal-most D4Z4 unit reported; (**B**) Plots of individual read D4Z4 copy number and distal-most D4Z4 unit methylation across diagnostic groups. Colours indicate allele haplotype (4qA, 4qB, 10qA) and read type (complete or partial). Horizontal dashed lines represent the 10-copy threshold for FSHD1 pathogenic contraction, and vertical dashed lines mark 50% methylation. Representative examples are shown for FSHD1 (case L150, with a distal-hypomethylated 5-copy D4Z4 array on a 4qA haplotype and normal distal methylation of the non-contracted 4qA allele), non-FSHD control (case L249, with a non-hypomethylated, non-contracted D4Z4 array on a 4qA haplotype), and FSHD2 (case L153, with no D4Z4 contraction but significant hypomethylation of all [4qA, 4qB & 10qB] alleles). (**C**) Individual read 4q D4Z4 copy number distribution across the cohort. The dashed horizontal line marks the 10-copy threshold used to define pathogenic FSHD1 contractions. (**D**) Limit of detection analysis for D4Z4 copy number. Each panel shows the probability of detecting reads completely spanning the D4Z4 array when simulating increased sample multiplexing in representative individuals carrying 5-, 6-, 10-, and 11-copy D4Z4 arrays. Curves correspond to detection thresholds requiring ≥1 read (red), ≥2 reads (green), or ≥3 reads (blue). Detection probability decreases with higher multiplexing factors due to reduced per-sample coverage. (**E**) Distribution of distal-most 4qA D4Z4 unit methylation across the cohort. Boxplots summarise per-read methylation fraction at the distal-most D4Z4 unit for an individual based on complete or partial distal 4qA reads. Colours distinguish read type (complete 4qA or partial distal 4qA). Background shading indicates sample groups: FSHD1 (pink), FSHD1+2 (purple), FSHD2 (blue), borderline FSHD1 with a 11-copy D4Z4 array (green), and non-FSHD cases (grey). Solid horizontal lines show the median, and dashed lines the median ± 2 × median absolute deviation. Blue reference lines indicate complete 4qA reads from FSHD1 cases, and orange reference lines indicate partial distal 4qA reads from non-FSHD cases.

We first evaluated our method via analysis of 10 cases diagnosed with FSHD1 by SB, 3 cases of FSHD2 (from a single family) and 1 case of FSHD1+2. We detected a pathogenic D4Z4 repeat contraction (≤10 repeats) in cis with a permissive 4qA haplotype in 9/10 FSHD1 cases (Table 2; Fig. 2C). The remaining case (L117) was shown not to harbour a contracted D4Z4 array on a 4qA allele (see below). The D4Z4 repeat number identified by **d4z4ling** matched the D4Z4 repeat number estimated by SB exactly in 4/10 cases and was within 2 repeats of all SB estimates (Table 2). The margin of error in the D4Z4 repeat number count estimated by SB increases progressively with increasing D4Z4 repeat size (reported by our local diagnostic laboratory as ± 2 kb for a 16.3 kb fragment and ± 4.2 kb for a 45.3 kb fragment). In contrast, **d4z4ling** provides a precise readout of the D4Z4 repeat number supported by multiple independent ‘complete’ sequencing reads. The observed discrepancies are therefore consistent with imprecision in SB size estimates. To assess minimum sequencing requirements to confidently detect D4Z4 contractions, we conducted a downsampling experiment using data from four cases with 5-, 6-, 10- and 11-copy D4Z4 alleles (see **Methods**). For all cases, complete D4Z4 (4qA) reads were detected with >90% confidence at a multiplexing rate of 3 samples per flow cell. Reducing this to 2 samples per flow cell was necessary to ensure 100% confidence for the 10- and 11-copy arrays (Fig. 2D & <u>S7</u>). While performance may vary based on the specifics of an individual case, these results indicate that reliable FSHD profiling can be achieved when running 2–3 samples per flow cell, thus establishing a streamlined and comprehensive strategy with sufficient data output for confident detection of D4Z4 repeat contractions.

In addition to D4Z4 contractions, FSHD1 cases showed the expected methylation profile with hypomethylation of the distal-most D4Z4 unit on the contracted 4qA D4Z4 repeat array compared to participants without FSHD (median ± median absolute deviation [MAD] methylation 29% ± 8.29% vs. 68% ± 7.58%, respectively) and normal methylation of the distal-most D4Z4 unit on the other, non-contracted allele (Fig. 2E). Comparing the median ± 2 × MAD of the distal D4Z4 unit between the two groups yields distinct, non-overlapping ranges: 12.42–45.58% for FSHD1 vs. 52.84–83.16% for non-FSHD (Fig. 2E). In the family of three patients with FSHD2, in addition to detecting a previously identified intronic *SMCHD1* variant (<u>Fig. S5A</u>) that had been shown to be pathogenic by an external research laboratory (data not presented), we observed distal D4Z4 unit hypomethylation on both 4q alleles (Fig.2E); median methylation fell within the previously identified median ± 2 × MAD range seen in FSHD1 patients. In the one case of FSHD1+2 (L439), our assay identified both the known 7-copy D4Z4 contraction on a 4qA haplotype and *SMCHD1* variant (<u>Fig. S5B</u>). Both 4q D4Z4 alleles were hypomethylated, similar to FSHD2 patients, but the contracted 4q D4Z4 allele was much more severely hypomethylated than the non-contracted allele (Fig. 2E & <u>S5C</u>), highlighting the compounding effects of the two pathogenic variants in this participant. Our data also recapitulates known relationships between D4Z4 copy number, D4Z4 methylation level and age of symptom onset in FSHD1 (<u>Fig. S6</u>).^5,21^

### Enhanced genetic and epigenetic characterisation of FSHD

Our new FSHD analysis method provided a more complete, high-resolution survey of FSHD loci than previously possible, resulting in several new, revoked and/or refined diagnoses and new genetic insights amongst our cohort.

In cases L246 and L247 (Fig. 3A), siblings with FSHD1, SB results not only identified two D4Z4 contraction lengths, these lengths were apparently discordant between siblings (7 & 9 D4Z4 repeats in L246; 6 & 8 D4Z4 repeats in L247). Additionally, results of previous BSS performed by an external research laboratory^22^ were also discordant, being reported as consistent with FSHD1 in L246 and FSHD2 in L247. LRS data resolved these discrepancies, showing that both siblings harboured biallelic, hypomethylated D4Z4 contractions of 6 and 8 repeats, both on 4qA alleles (Fig. 3B-C). The uncommon occurrence of biallelic 4qA contractions, presumably resulting in lower methylation levels than typically seen in FSHD1, is the likely reason that the previous BSS assay reported a diagnosis of ‘FSHD2’ in L247 and highlights the added utility of our assay over BSS (and SB) in these complex cases.

**Figure 3.**
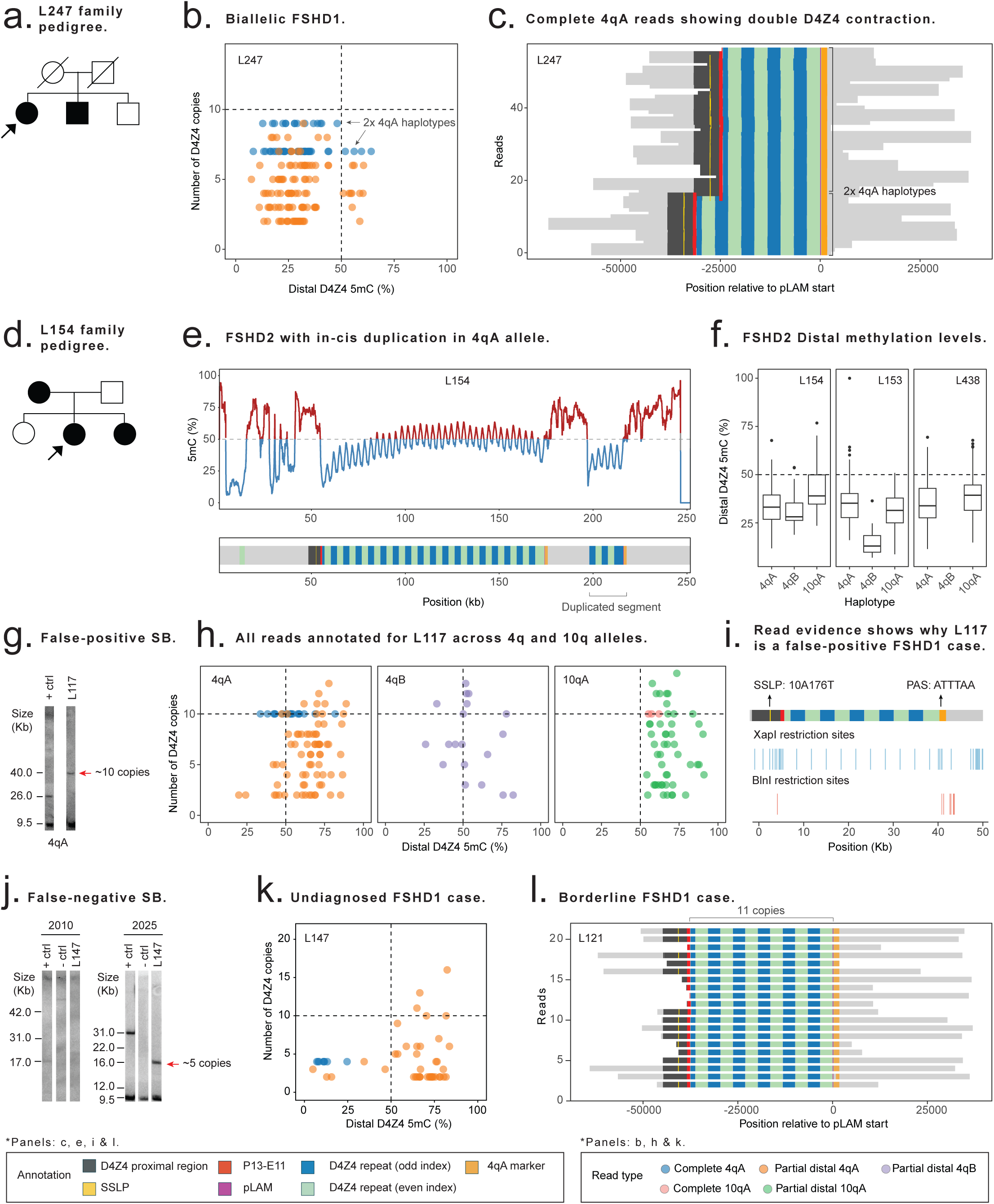
Targeted ONT LRS clarifies diagnosis in complex FSHD cases. (**A**) Pedigree of L246 and L247, siblings with FSHD1. Squares represent males and circles females; filled symbols indicate affected individuals. The proband (L247) is indicated with an arrow. (**B**) Plot of individual read 4qA D4Z4 copy number and 4qA distal-most D4Z4 unit methylation in case L247 demonstrate the presence of both a 9- and a 7-copy D4Z4 contraction allele, both hypomethylated. Colours reflect haplotype (4qA) and read type (complete or partial). The horizontal dashed line marks the 10-copy threshold for pathogenic FSHD1 contractions, and the vertical dashed line indicates 50% methylation. (**C**) Read pile-up of complete 4qA reads from case L247, again showing biallelic D4Z4 contractions. Each horizontal bar represents a single long read aligned relative to the pLAM start position, with annotated features across each read. Two distinct groups of reads are observed, corresponding to contractions of 7 and 9 D4Z4 repeat units, consistent with biallelic contraction. (**D)** Pedigree of L438, L153 and L154, a mother and two daughters with FSHD2. Symbols as in panel A. The proband (L154) is indicated with an arrow. (**E)** Methylation profile across the 4qA allele in case showing an in *cis* duplication of the D4Z4 array which contains 5 D4Z4 units and is hypomethylated. The top panel shows CpG methylation fraction (5mC %) across the locus; red indicates >50% methylation and blue indicates <50%, with the dashed line marking 50%. The bottom schematic shows the assembled 4qA allele generated with Hifiasm, against which individual reads were mapped. (**F)** Distal-most D4Z4 unit methylation levels in L153, L154 and L438. Each boxplot summarises per-read methylation fraction at the distal-most D4Z4 unit, stratified by haplotype (10qA, 4qA, 4qB). The dashed horizontal line indicates 50% methylation. (**G)** Historical 4qA-specific Southern blot gel of case L117, showing a band at 40 kb (∼10 D4Z4 units). (**H)** Plot of individual read D4Z4 copy number and distal-most D4Z4 unit methylation in case L117 across all alleles (4qA, 4qB, 10qA) showing the 10-copy D4Z4 contraction incorrectly classified as 4qA, with reads mapping to both 4q and 10q D4Z4 loci. Plot layout and annotations as in panel B. (**I)** Representative annotated contracted LRS reads from case L117 with D4Z4 features, haplotype markers (SSLP: 10A176T, PAS: non-canonical ATTAA), and restriction enzyme recognition sites (*XapI* and *BlnI*). This analysis confirms the 10-copy D4Z4 contraction was not on a 4qA allele but on an uncommon 10qA allele, 10A176T, resulting in the false positive Southern blot. (**J)** Historical linear gel electrophoresis Southern blot of case L147 showed no D4Z4 contraction. Following ONT LRS diagnosis, repeat Southern blot following pulsed-field gel electrophoresis confirmed the presence of a contracted 4qA band at 16 kb (∼5 D4Z4 units). (**K)** Plot of individual read 4qA D4Z4 copy number and distal-most 4qA D4Z4 unit methylation in case L147 demonstrates a distally hypomethylated 4-copy D4Z4 array. Plot layout and annotations as in panel B. **(L)** Read pile-up of complete 4qA reads from case L147, showing a 11-copy D4Z4 repeats. Annotations as in panel C.

In cases L153, L154, and L438, two daughters and a mother with FSHD2 (Fig. 3D), in addition to detecting the known *SMCHD1* variant and FSHD2 methylation pattern, LRS identified that all three also harboured permissive 4qA alleles containing a duplicated D4Z4 repeat array, with at least one duplicated D4Z4 segment that was 5-copies in length and hypomethylated (Fig. 3E-F). Such in *cis* D4Z4 array duplications have been reported previously in FSHD with the size of the individual duplicated D4Z4 arrays thought to determine whether the duplication by itself causes autosomal dominant FSHD1 or whether an additional variant in *SMCHD1* is required, causing digenic FSHD2.^23^ Using an empiric formula determined by Lemmers *et al.*^24^, the in *cis* duplication event in the family studied here is predicted to require an additional FSHD2 variant to cause FSHD.

Our analysis also clarified several cases where SB results either led to incorrect diagnoses or were inconclusive. Case L117 previously had a SB showing a 10-copy D4Z4 contraction apparently located on a permissive 4qA allele (Fig. 3G). However, our LRS data unexpectedly showed this participant did not have a D4Z4 contraction on a 4qA allele nor distal D4Z4 hypomethylation (Fig. 2E), but rather a 10-copy D4Z4 contraction on a rare 10qA haplotype known as 10A176T (Fig. 3H-I). The 10A176T haplotype, present in 2.5% of the general European population, contains chromosome 4-like restriction enzymes sites and is theorised to be derived from an ancestral chromosome 4 to chromosome 10 translocation event.^25,26^ The presence of a D4Z4 contraction on this 10A176T allele appears to have resulted in a false-positive SB finding in case L117. Although an alternative genetic myopathy diagnosis was not identified, our results clearly indicate this participant does not have FSHD1 or FSHD2 and subsequent review of the clinical phenotype revealed the presence of clinical features atypical for FSHD (symmetric weakness, presence of brisk reflexes).

Case L147, recruited as an unsolved Group 2 case, had a pattern of weakness consistent with FSHD but SB (following linear gel electrophoresis; LGE) had previously shown the absence of a D4Z4 repeat contraction (Fig. 3J). Our LRS assay showed this participant harboured a hypomethylated 4-copy D4Z4 contraction on a permissive 4qA allele, consistent with FSHD1 (Fig. 3K). Subsequently, repeat SB (following pulsed-field gel electrophoresis; PFGE) was performed, confirming a contraction of 16 kb (∼4 D4Z4 repeats) (Fig. 3J). The cause for the initial negative SB likely relates to technical limitations of LGE-SB compared to PFGE-SB. As a result of the new diagnosis, case L147 and his family could be offered genetic counseling specific to the autosomal dominant inheritance pattern of the disease, which was not previously apparent as both parents were unaffected, and referral for participation in specific FSHD1 clinical trials which require a secure genetic diagnosis for enrolment.

Case L121 had a typical FSHD phenotype but a previous SB showed a 4qA allele with an estimated 11 D4Z4 units, which is not thought to cause FSHD1. Given the known error margin for SB D4Z4 array sizing, it was suspected that the participant might actually have a 10-copy D4Z4 array and the patient was considered to have ‘possible FSHD1’. However, our LRS data confirmed an 11-copy D4Z4 array on a permissive 4qA haplotype, supporting the original SB findings (Fig. 3L). The distal-most D4Z4 unit of the 11-copy 4qA D4Z4 array was hypomethylated (Fig. 2E), while the other 4q allele (which was a 4qB haplotype) was not hypomethylated. This pattern and the absence of any disease-causing variants in *SMCHD1*, *DNMT3B* and *LRIF1* argued against a diagnosis of FSHD2. No alternative genetic explanation for the participant’s FSHD-like phenotype was identified. While this participant remains genetically undiagnosed, our findings raise the question of whether 11-copy D4Z4 repeat 4qA alleles, which are imprecisely measured using SB and currently considered non-pathogenic, may occasionally cause FSHD1 (contrary to current thinking), perhaps with incomplete penetrance. LRS analysis and careful phenotyping of further cases with 11-copy D4Z4 repeats is required to investigate this further.

### Enhanced genetic and epigenetic characterisation of OPDMs

Out of the 9 new diagnoses established by our LRS assay, 7 were OPDM, a group of disorders caused by non-coding GGC•CCG STR expansions, typically located in the 5’ untranslated region. Four patients were diagnosed with *GIPC1*-related OPDM (OPDM2) and one each with *NOTCH2NLC*-related (OPDM3), *RILPL1*-related (OPDM4) and *ABCD3*-related OPDM (Table 3; Fig. 4). Skin biopsies were subsequently performed in 4 of these cases, and all showed the presence of p62- and ubiquitin-positive intranuclear inclusions (INIs), typical of OPDM (Fig. 4E; **Supplementary Methods**). Our results demonstrate that OPDM appears to be a major contributor to currently undiagnosed genetic myopathies.

**Figure 4.**
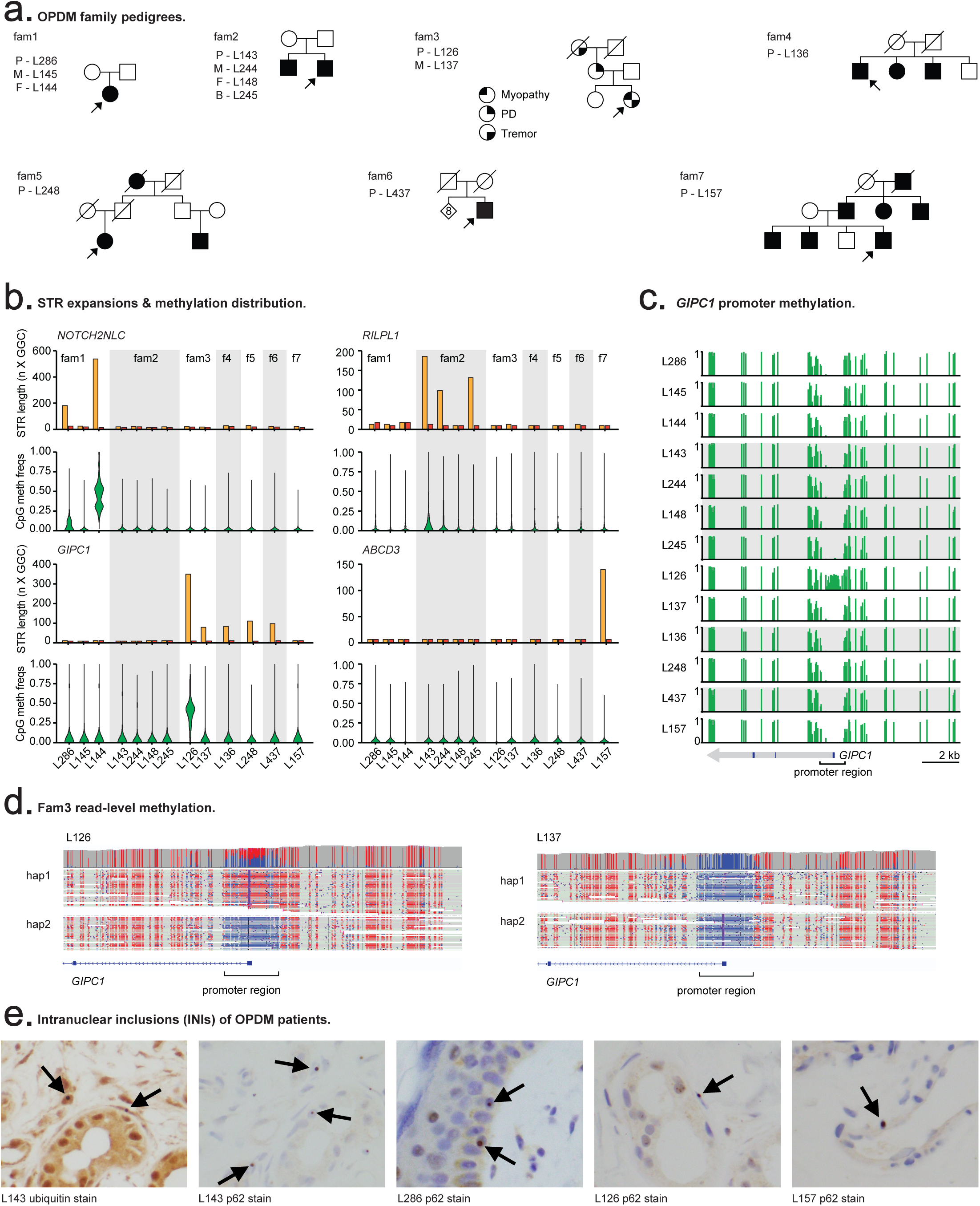
Targeted ONT LRS characterises the genetic and epigenetic architecture of OPDM. (**A**) Pedigrees of OPDM families diagnosed by ONT LRS (fam1–fam7). Squares denote males and circles females; filled symbols indicate affected individuals. The proband is marked with an arrow, and IDs for proband (P), mother (M), father (F) and brother (B) are shown for family members who underwent LRS. **(B)** GGC•CCG STR expansions and CpG methylation distribution across individuals in OPDM families; . For each locus (*NOTCH2NLC*, *RILPL1*, *GIPC1*, and *ABCD3*), the top panels show allele-specific STR repeat lengths (yellow = haplotype 1, orange = haplotype 2). Bottom panels show per-read CpG methylation distributions, with violin plots representing the density of methylation values. Family members are grouped by shaded backgrounds. **(C)** CpG methylation across the *GIPC1* promoter in OPDM families. Vertical bars indicate methylation levels at different CpG sites. L126 is the only individual demonstrating *GIPC1* promoter hypermethylation. **(D)** Genome browser view of *GIPC1* read-level methylation in L126 and L137; reads are phased into haplotypes 1 and 2. CpG sites are coloured by methylation state (red = methylated, blue = unmethylated). In L126, a participant with OPDM2 due to a *GIPC1* (GGC)217-570 STR expansion, allele-specific hypermethylation is evident across the large expanded allele and the *GIPC1* promoter region. In contrast, the promoter is unmethylated in the participant’s mother (L137) who has Parkinson’s disease but no myopathy and a *GIPC1* (GGC)76-85 STR expansion. **(E)** Anti-ubiquitin and anti-p62 immunohistochemistry of skin biopsies (×400 magnification) identifies intranuclear inclusions in a range of cell types (epithelium, adipocytes, fibroblasts, pericytes) in OPDM cases.

Case L286 (fam1 in Fig. 4A) had a GGC expansion in *NOTCH2NLC* of variable length (range 130–621 repeats) and skin biopsy showed numerous p62/ubiquitin-positive INIs (Fig. 4E), consistent with a diagnosis of OPDM3. The participant’s father (L144), who did not have myopathy, was found to harbour a very large GGC expansion, again with variable length (range 273–905). The variable length of the *NOTCH2NLC* STR expansions are consistent with somatic instability/mosaicism that has been described in some patients.^27^ While the STR expansion and the surrounding *NOTCH2NLC* promoter region in L286 exhibited DNA methylation levels similar to non-OPDM3 participants and healthy controls, LRS identified that the same region in L144 exhibited allele-specific hypermethylation of the *NOTCH2NLC* promoter region (Fig. 4B & <u>S8</u>). This phenomena of asymptomatic parents harbouring extremely long ([GGC]_>400_), hypermethylated expansions in *NOTCH2NLC* having children with shorter, but still pathological, GGC expansions that are not hypermethylated, is well documented in *NOTCH2NLC*-related OPDM3 and neuronal intranuclear inclusion disease (NIID).^27,28^ Contraction of the GGC STR expansion size across generations occurs more commonly following paternal inheritance,^27^ as was the case here. Case L286 and her asymptomatic father both also harboured a heterozygous expansion of the normal 29–31 × 99 bp VNTR in exon 5 of *PLIN4* to 39 × 99 bp (<u>Fig. S9</u>). Although *PLIN4* VNTR expansions have been described to cause a vacuolar myopathy, often with prominent distal weakness,^29^ the significance of the VNTR expansion in this family is unclear given that OPDM3 better accounts for the phenotype and that the father is asymptomatic (<u>Supplementary Note 2</u>).

Cases L126, L136, L248 and L437 (fam3, fam4, fam5 and fam6 in Fig. 4A, respectively) were found to have GGC STR expansions (217–570, 85, 112 and 99 repeats, respectively) in *GIPC1*. The previously reported pathogenic range for *GIPC1* GGC expansions was 73–164, indicating that cases L136, L248 and L437 can be confidently diagnosed with OPDM2.^30^ Similar to *NOTCH2NLC*, extremely long, hypermethylated STR expansions in *GIPC1* ([GGC]_>500_) have been found in asymptomatic fathers of OPDM2 patients who themselves have typical *GIPC1* repeat expansion lengths.^30^ We observed allele-specific DNA hypermethylation of the large variable-length (GGC)_217–570_ expansion and surrounding *GIPC1* promoter region in L126, compared to all other samples (Fig. 4B-D). Based on the clinical presentation and presence of INIs on skin biopsy (Fig. 4E), we diagnosed case L126 with OPDM2. This would seem to expand the pathogenic STR length for *GIPC1* to at least 217 repeats and demonstrates that OPDM may still manifest in cases of hypermethylated GGC repeats. The mother of case L126 (L137), did not have myopathy, but did have typical, levodopa-responsive Parkinson’s disease. She (L137) was found to harbour a non-hypermethylated (GGC)_76–85_ expansion in *GIPC1* (Fig. 4B) but skin biopsy did not identify any INIs. This highlights the instability of the *GIPC1* repeat across generations and also aligns with recent reports linking shorter *GIPC1* STR expansions with Parkinson’s disease.^31,32^ The *GIPC1* STR expansions in cases L248 (New Zealand Māori ethnicity) and L437 (mixed New Zealand Māori, Tongan and European Australian ethnicity) occur within a haplotype consisting of 7 SNPs in or near the *GIPC1* locus, that was previously identified in 100% of a Japanese cohort of 10 OPDM2 patients but which is only present in 7% of non-expanded Japanese *GIPC1* alleles and 11% of the general population.^33^ In contrast, cases L126 (European Australian) and L136 (Chinese) only harboured 6 of the 7 SNPs on their respective *GIPC1* STR expansion alleles, suggesting that, compared to the presumed New Zealand Māori *GIPC1* allele, these cases may have a more distant shared ancestral origin with Japanese OPDM2 cases (<u>Table S2</u>).

Case L143 (fam2 in Fig. 4A) was found to have a (GGC)_191_ expansion in *RILPL1* consistent with OPDM4 and skin biopsy identified the typical INIs (Fig. 4E). His brother (L245), who had ocular and facial weakness without limb or pharyngeal involvement and had been labelled as having chronic progressive external ophthalmoplegia (CPEO), a type of mitochondrial myopathy, was recruited to this study. LRS showed the brother harboured a (GGC)_131_ expansion in *RILPL1* and no pathogenic variants in any of the nuclear or mtDNA genes that cause CPEO, indicating he also has OPDM4. Their mother (L244), who did not have myopathy or any other neurologic features, was found to harbour a (GGC)_98_ expansion in *RILPL1*, falling short of the previously reported pathogenic size range of 130–197.^34^ The mother’s STR allele represents an ‘intermediate range’ expansion that does not cause symptoms in itself but is prone to further expansion in subsequent generations, as seen in this family. The *RILPL1* STR and promoter regions were not hypermethylated in any of these individuals.

Case L157 (fam7 in Fig. 4A), only presented with ptosis and subtle ophthalmoplegia but had a family history of an autosomal dominant OPDM phenotype. He was identified to have a (CCG)_170_ expansion in *ABCD3* (Fig. 4B) and was subsequently identified to be related to one of the recently published European Australian *ABCD3*-OPDM families.^35^

### Identification of a *LAMA2* splice-altering variant

Case L236 had a limb girdle pattern of myopathy and an apparently autosomal dominant pattern of inheritance (Fig. 5A). A previous NGS targeted gene panel (which included *LAMA2*) did not identify any causative genetic variants and the case remained genetically unsolved. Our LRS assay identified a homozygous intronic variant in *LAMA2*, NM_000426.4:c.2749+4_2749+15del, affecting the extended donor splice site of intron 19 (Fig. 5B). Sanger sequencing confirmed the homozygous status of the variant in the proband and that the proband’s mother was a heterozygous carrier (Fig. 5C). The proband’s affected father and paternal uncle were deceased and, thus, unable to be tested. The variant (rs2482293091) is present in one South Asian individual on gnomAD v4.1.0 and has previously been reported as a VUS in ClinVar (RCV003133994.3 and RCV004577033.1). It was predicted by SpliceAI^36^ to result in loss of the donor site (Δ0.99) and use of a cryptic donor splice site 78 bp upstream (Δ0.55). Targeted RT-PCR from participant skin-derived fibroblasts (**Supplementary Methods**) confirmed that the normal *LAMA2* splicing product was absent and demonstrated the presence of two aberrant splicing products. The predominant product resulted from use of the cryptic upstream donor site resulting in an in-frame 75 bp deletion-insertion, p.Gly892_Pro917delinsAla (Fig. 5D). This same mis-splicing event, alternatively labelled as p.Glu892Ala del893_917, was previously identified in a patient with a different variant affecting the same donor splice site (c.2750+2insT).^37^ RT-PCR also identified that complete skipping of exon 19 was another, less frequent, mis-splicing event (Fig. 5D). As a result of the splicing studies, we considered this intronic *LAMA2* variant to be pathogenic and diagnosed case L236 with *LAMA2*-related limb-girdle muscular dystrophy (LGMD) type R23. Review of a previous muscle MRI and subsequent brain MRI identified features consistent with LGMD R23 (Fig. 5E-F).^37^

**Figure 5.**
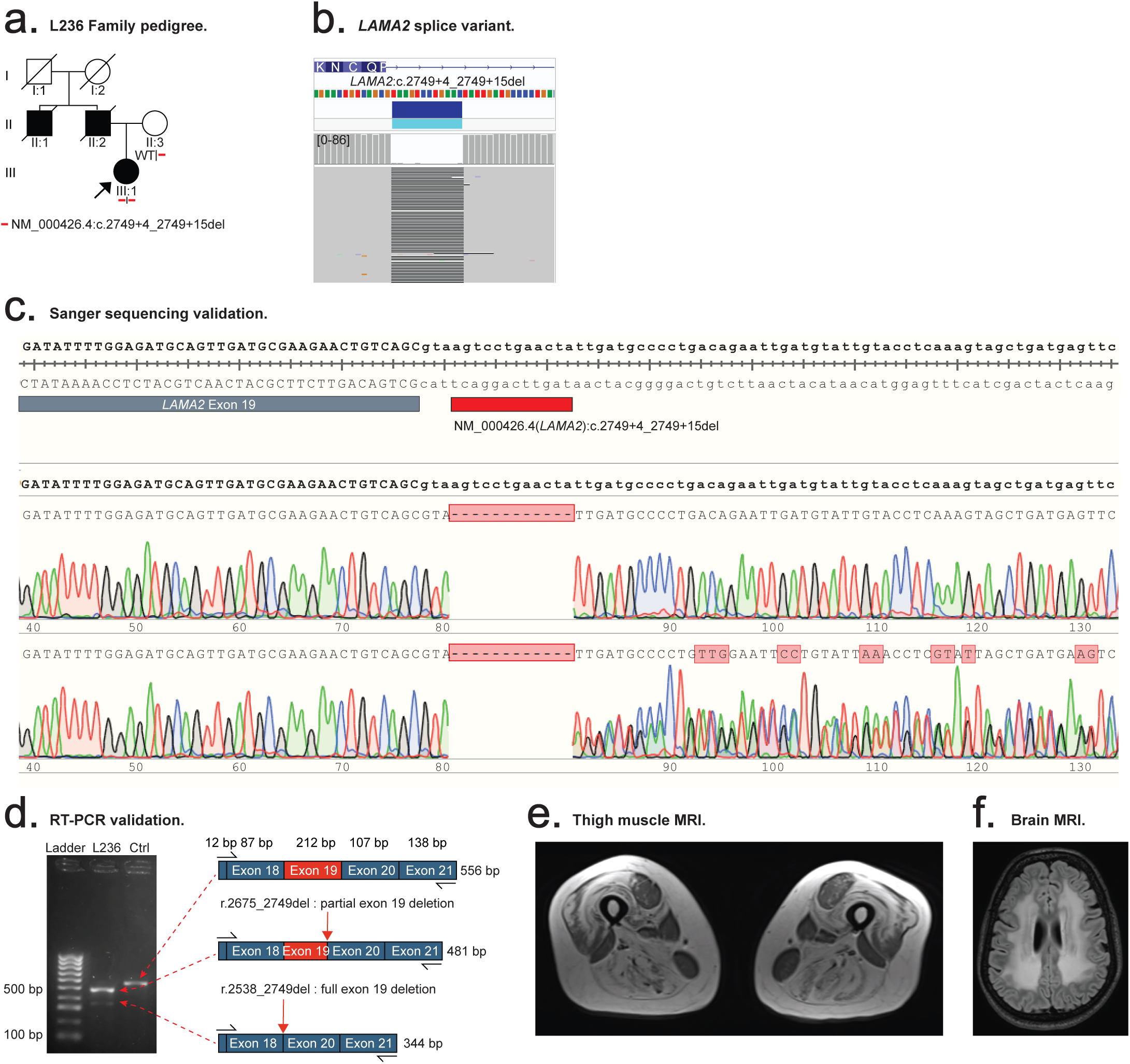
Targeted ONT LRS lead to a new diagnosis of LGMD R23. (**A**) Pedigree of case L236. Squares represent males and circles females; filled symbols indicate affected individuals. The proband (L236) is indicated with an arrow. **(B)** Genome browser view of a homozygous intronic variant in *LAMA2* (c.2749+4_2749+15del) identified by ONT LRS. **(C)** Sanger sequencing validation of the *LAMA2* variant. Chromatograms from the proband (top track) and proband’s mother (bottom track) confirm the proband is homozygous and the mother is heterozygous for the variant (indicated by apparent frameshift following the deletion). **(D)** RT-PCR validation of the *LAMA2* splice variant. Gel electrophoresis of *LAMA2* RT-PCR products (distal exon 17 to exon 21) from skin biopsy-derived fibroblasts in the L236 and a control (Ctrl) show an absence of the normal splice product in L236 and the presence of two aberrant transcripts corresponding to partial and complete exon 19 deletion. **(E)** Thigh MRI showing a pattern of muscle involvement consistent with LGMD R23, with relative sparing of the rectus femoris, gracilis, sartorius, and short head of the biceps femoris. **(F)** Brain MRI showing confluent white-matter T2 hyperintensity, characteristic of LGMD R23.

The apparent dominant inheritance in this family was presumed to reflect pseudodominance and the homozygosity of the variant in L236, in the absence of consanguinity, attributed to identity by descent – both parents’ families came from the same small city in India. Pseudodominance has previously been described in *LAMA2*-related muscular dystrophy.^37^ As a result of the new diagnosis, the patient and her family were provided with genetic counselling pertaining to autosomal recessive inheritance, overriding the previous erroneous assumption of autosomal dominant inheritance.

## Discussion

Our targeted ONT LRS assay and analysis framework provides thorough characterisation of the wide range of genetic and epigenetic variation that cause genetic myopathies. We identified a new genetic diagnosis in almost a third of patients who had undergone previous standard genetic testing. Our assay also improved the precision of previously known genetic diagnoses and also identified several false-positive and false-negative diagnoses that were overturned using LRS data – errors that highlight the clinical and analytical challenges involved. The new diagnoses achieved in this study resolved prolonged diagnostic odysseys, spanning years to decades (mean disease duration of 14 years for the n = 9 newly solved cases) and facilitated disease-specific management (e.g. genetic counselling, clinical trial participation, etc.) In addition to improved diagnosis, the high resolution and comprehensive nature of LRS data also shed new light on the molecular basis of disease. For example, our FSHD analysis method observed the compounding effects of ‘double’ variants in a case of FSHD1+2. Similarly, our results help to refine the observed pathogenic range for STR expansions in patients with OPDM and characterise the influence of DNA methylation in this disorder.

We showed that LRS is capable of comprehensively assessing the complex genetic and epigenetic architecture of FSHD. Our **d4z4ling** software package provides accurate D4Z4 repeat copy number assessment, single-molecule methylation profiling, and detailed haplotype resolution. While ONT-based methods have previously been applied to FSHD1,^38–41^ our approach is the first to demonstrate the ability of adaptive targeted LRS to characterise FSHD1, FSHD2, FSHD1+2 and all other myopathic loci concurrently, simplifying patient analysis in the face of genotypic and phenotypic heterogeneity. **D4z4ling** is compatible with any long-read data type and we provide it as open source software for the genomics community.

OPDM accounted for >75% of new diagnoses identified by our LRS assay, which is perhaps unsurprising given the current lack of clinically accredited genetic testing options for any of the OPDM STR expansions in Australia. The OPDM phenotype was described in the 1970s but the causative GGC•CGG STR expansions were only identified in the last 6 years.^30,34,35,42,43^ These STR expansions were initially described predominantly in East Asian cohorts, whereas the most recently identified locus (*ABCD3*) was found in OPDM patients of European ancestry.^35^ Our LRS assay was not only able to concurrently identify the presence of STR expansion across multiple loci but also characterise the methylation status of these loci, characterise mosaicism of the STR expansion length, and phase the STR expansions with surrounding SNPs to identify common haplotypes. Our results expand knowledge of the ethnic backgrounds in which different OPDM types occur. The finding of a *NOTCH2NLC* GGC expansion causing OPDM3 that originated from a Cook Island Māori heritage extends the recent description of *NOTCH2NLC* GGC expansion-related NIID in this ethnic population.^44^ To date, *GIPC1*-related OPDM2 has only been reported in East Asian patients. For the first time, we report *GIPC1* OPDM2 in patients of European Australian and New Zealand Māori heritage. The sharing of uncommon SNPs with a Japanese OPDM2 haplotype suggests the possibility of a common ancestral haplotype that may predispose to expansion of the *GIPC1* STR. *RILPL1* GGC expansions, which cause OPDM4, have only ever been described in OPDM patients of Chinese heritage and were even absent in a large Japanese OPDM cohort.^45^ Here, we report the novel finding of OPDM4 in a family of mixed Papua New Guinean and Ni-Vanuatu ethnicity. An OPDM phenotype has previously been reported in a set of Papua New Guinean siblings, many years prior to identification of the causative STR expansions;^46^ in light of our findings, it is interesting to speculate whether that family may have had OPDM4. Similarly, it would be of interest to determine if ‘Simbu ptosis’, a familial disorder apparently endemic to the Eastern Highlands of Papua New Guinea, consisting of myogenic ptosis ± ophthalmoplegia, facial weakness, nasal voice and foot drop (all features of OPDM),^47^ also represents OPDM4.

Despite our approach having clear benefits in terms of improved molecular characterisation of genetic myopathies, our study has several limitations. The use of adaptive targeted ONT sequencing, compared to whole-genome LRS (ONT or PacBio), represents a more cost-effective assay on a per participant basis, but is restricted to the programmed target loci, preventing analysis of new myopathy genes discovered subsequent to the time of sequencing. Fortunately, unlike traditional targeted NGS panels, which require redesign of molecular probes or primers used for target enrichment, the *in silico* panel used to guide our ONT adaptive sampling assay can be updated simply by adding new genomic coordinates to a digital file. For example, we were able to rapidly add in the *ABCD3* loci during the course of our study following its identification as a cause for OPDM in 2024.^35^ Next, pre-study diagnostic workup varied widely between participants in Group 2 of our cohort (e.g. some had a targeted NGS panel, others had prior exome or genome NGS, etc.), making it difficult to precisely quantify the added diagnostic yield of ONT LRS on top of each of these individual NGS assays. However, this situation reflects the heterogeneity of a ‘real world’ cohort of unsolved genetic myopathy patients and here we have shown that ONT LRS increases overall diagnostic yield and accuracy compared to the collective group of ‘standard’ genetic testing methodologies.

In conclusion, we describe a new targeted ONT LRS assay capable of assessing the full range of genetic and epigenetic variants implicated in the genetic myopathies and demonstrate its utility for diagnosis of patients with these conditions. Our method provides deep insights into the complexity of this heterogeneous group of disorders, resolving challenging cases and revising erroneous diagnoses made by older molecular techniques. Our results demonstrate that clinical adoption of LRS will simplify diagnostic algorithms, improve diagnostic rates, and advance scientific understanding of the genetic myopathies.

## Materials & Methods

### Study Participants

Participants were recruited from patients referred to Concord Repatriation General Hospital and Neuroscience Research Australia from March 2023 to September 2025. A positive control cohort, Group 1, consisted of patients with genetically confirmed myopathies specifically selected to represent a range of different genetic and epigenetic variant types. An undiagnosed cohort, Group 2, consisted of consecutively recruited participants that met the following criteria: 1) history, examination and investigations (creatine kinase, neurophysiology, muscle imaging and/or muscle biopsy) suggestive of and concordant for a myopathic process; 2) myopathy was the predominant neurologic phenotype; 3) the treating/referring clinician assessed that a genetic cause for the myopathy was more likely that an acquired aetiology; and 4) the participant remained genetically unsolved (or incompletely solved) after appropriate conventional clinical genetic testing as determined by the treating/referring clinician. Where relevant, family members of probands were recruited for segregation studies. The protocol for this study was approved by the St Vincent’s Hospital Human Research Ethics Committee (2019/ETH12538) and all participants provided written informed consent. **Patient IDs mentioned are not known to anyone outside the research team**. Pedigrees have been simplified or modified to protect the anonymity of families.

### Targeted ONT LRS

Peripheral blood samples were obtained and high-molecular weight (HMW) genomic DNA was extracted using the Nanobind CBB kit (PacBio, Cat# 102-301-900) or PanDNA kit (PacBio, Cat# 102-260-000).

HMW genomic DNA was sheared to ∼40–50 kb fragment size using the Diagenode Megaruptor 3 DNA shearing system (speed 27), treated with the Short-read Eliminator kit (PacBio, Cat# 102-208-300) to deplete fragments <10 kb, then visualised on an Agilent Femto Pulse using the Genomic DNA 165 kb Kit. ONT libraries were prepared from ∼4 μg of sheared HMW genomic DNA using a ligation prep (SQK-NBD114.24). Between 1–3 participant samples were barcoded and pooled into one library and loaded on an ONT PromethION R10.4.1 flow cell (FLO-PRO114M) and sequenced on either a PromethION 2 Solo or PromethION 48 instrument.

Readfish^48^ (v0.0.10dev2) was used for live target selection or rejection utilising a custom panel of 331 loci associated with genetic myopathies (<u>Table S1</u>). These loci were chosen based on a literature review performed in January 2023, focusing on genetic myopathic disorders, supplemented by review of existing myopathy gene panels in PanelApp and from diagnostic genomic laboratories (Invitae, Blueprint Genetics and PathWest). The *ABCD3* locus was added to the panel during the course of this study, such that it was sequenced in 26 out of the 53 proband samples. Gene targets were encoded relative to the T2T-chm13 (v2) reference genome and ReadFish was executed with the following configurations: config_name = ‘dna_r10.4.1_e8.2_400bps_5khz_fast_prom’; min_chunks = 0; max_chunks = 16; single_on = ‘stop_receiving’; multi_on = ‘stop_receiving’; single_off = ‘unblock’; multi_off = ‘unblock’; no_seq = ‘proceed’; no_map = ‘proceed’. Libraries were run for 72 hours, with nuclease flushes and library reloading performed at approximately 24-hour and 48-hour time points.

### LRS Data Analysis

After completing a targeted sequencing run, raw ONT sequencing data were converted to BLOW5 format^49^ and base-called with Buttery-eel^50^ (a wrapper to enable BLOW5 input for ONT’s Dorado basecaller), using the ‘super-accuracy’ model with 5mC methylation calling enabled. Base-called data was then processed using pipeface (https://github.com/leahkemp/pipeface), a NextFlow pipeline for LRS data analysis. Pipeface executed minimap2^51^ (v2.28-r1209) for alignment, Clair3^52^ (v1.0.9) or Deepvariant (v1.80) for calling small variants, sniffles2^53^ (v2.6.0) and cuteSV^54^ (v2.1.1) for calling SVs, WhatsHap^55^ (v2.3) for phasing, and minimod^56^ (v0.3.0) for DNA methylation profiling. Small variants and SVs were functionally annotated using variant effect predictor (VEP)^57^ (v112).

Variants were prioritised for further consideration based on the following criteria: (i) those annotated in ClinVar with a clinical significance of ‘pathogenic’, ‘likely pathogenic’ or ‘uncertain’; (ii) variants predicted to cause a frameshift; (iii) missense variants with a REVEL score > 0.5 and classified as ‘pathogenic’, ‘likely pathogenic’, ‘uncertain’ or without an assigned clinical significance; (iv) variants with high or moderate impact that are similarly classified as ‘pathogenic’, ‘likely pathogenic’, ‘uncertain’ or lacking an assigned clinical significance in ClinVar and (v) variants predicted to alter splicing, defined as having a SpliceAI delta score > 0.2. All pre-selected variants were within MANE Select transcripts and either were not found in gnomADv4 or had allele frequency of less than 0.01, resulting in a list of rare and potentially causative variants. Additionally, SVs intersecting protein-coding genes were prioritised for further consideration.

STR genotyping was performed by LongTR^58^ (v1.0), using a set of pre-defined STR sites known to be implicated in genetic myopathies (<u>Table S1</u>). All STR sites were also manually inspected in the IGV genome browser to ensure the validity of expansions detected and confirm none were missed. The *PLIN4* exon 5 VNTR was also genotyped using LongTR, and the resulting VCFs were queried (bcftools query -f ’[%SAMPLE\t%GB\n]’) to extract the GB field, which reports base-pair differences from the reference allele. Samples showing a marked deviation from the hg38 reference were reviewed in IGV to confirm the presence and boundaries of the insertion. To assess the potential protein impact, an alternative MANE Select transcript incorporating the expanded VNTR was generated and translated using ExPASy.^59^

Phased 5mC methylation frequency tracks produced by minimod were used to assess allele-specific DNA methylation at relevant gene loci.

Mitochondrial variants were called and annotated using the Mitoverse mtDNA-server 2^60^ with a detection limit of 0.1 variant allele fraction (VAF), with heteroplasmy level inferred from the VAF.

### FSHD Analysis

We developed a purpose-built, open source software package, **d4z4ling** (https://github.com/neysa-15/d4z4ling), to identify and characterise ONT reads selectively enriched with ReadFish^48^ targeting the D4Z4 region on chromosomes 4 and 10, using the T2T-chm13 (v2) reference genome (<u>Fig. S10</u>). Reads overlapping the D4Z4 loci were extracted and annotated individually for key features, including the proximal region, short sequence length polymorphism (SSLP), p13-E11 (5′ marker), pLAM (3′ marker), polyadenylation signal (PAS), *BlnI* and *XapI* restriction enzyme recognition sites and 4qA/4qB-specific probes. Reads were classified as ‘complete’, ‘partial proximal’, ‘partial internal’ or ‘partial distal’ depending on the extent to which they spanned the array. To estimate the number of D4Z4 repeat units, each read was aligned to a single D4Z4 repeat unit and the number of aligned copies was counted. For samples with complete reads and resolved copy numbers for one or both alleles, reads were phased based on D4Z4 repeat count and either polished into a consensus, or assembled using Hifiasm.^61^ Haplotypes were inferred based on SNVs and indels in the proximal region, SSLP length, sequence of the distal D4Z4 copy and presence of 4qA/4qB-specific features. CpG methylation levels were profiled at the distal D4Z4 copy with minimod, enabling integration of structural and epigenetic information at single-molecule resolution to inform FSHD diagnosis. Methylation status of each allele was assessed by assessing median and median absolute deviation of CpG methylation of the distal-most D4Z4 copy. Hypomethylation of the distal D4Z4 unit of a contracted 4qA D4Z4 array was considered indicative of FSHD1. Hypomethylation of the distal D4Z4 unit on both (non-contracted 4q) alleles was considered indicative of FSHD2, as long as one allele had a 4qA haplotype (i.e. either 4qA/4qA or 4qA/4qB).

Confident molecular diagnosis of FSHD1 requires complete reads spanning the full D4Z4 repeat array, enabling both precise repeat copy number estimation and methylation profiling. While ONT offers sufficiently long reads for this purpose, the likelihood of capturing a complete read decreases as the number of D4Z4 repeat units increases. To characterise this limitation, we used four samples (with pathogenic alleles carrying 5, 6, 10 and 11 D4Z4 units; <u>Fig. S7</u>) in which both 4q alleles could be resolved by either polishing ‘complete’ reads or *de novo* assembly. We progressively subsampled the read set to simulate decreasing sequencing depths. At each depth, we performed 50 random subsampling runs and recorded how often at least one complete read was detected. This allowed us to estimate the probability of detecting a complete read as a function of sequencing depth.

## Data Availability

Human participant data may be shared under restricted access to protect participant privacy. Enquiries for anonymised data access should be addressed to the corresponding authors K.R.K. and I.W.D.

## Acknowledgements

This study was supported by Medical Research Future Fund (MRFF) grants 2023126, 2025138 and National Health and Medical Research Council (NHMRC) grant 2035037. D.Y. is supported by a PhD Scholarship from Muscular Dystrophy NSW. K.R.K. is supported by the Ainsworth 4 Foundation and the MRFF. This project was undertaken with the assistance of resources and services from the National Computational Infrastructure (NCI), which is supported by the Australian Government and the Australian National University (ANU). The views expressed herein are those of the authors and are not necessarily those of the Australian Government, NHMRC or MRFF.

## Author Contributions

D.Y., K.R.K. and I.W.D. were involved in the conception of the study. D.Y., L.I.R., J.S.S., C.L., K.M., A.H., E.S., K.E.A., S.E., S.W.R., R.B., R.G., S.B., J.S., A.W., S.H., N.G.S., L.W., D.M., M.T., N.C.G., C.M.S., P.A.M., K.N., P.L.C. and K.R.K. were involved in clinical phenotyping. D.Y., A.L.M.R., I.S., N.N., L.K.., B.R.G., S.R.C., M.C., M.S., D.Z., R.L.D., M.L.K., P.L.C., K.R.K. and I.W.D. were involved in data generation, analysis and/or interpretation. D.Y. and A.L.M.R. drafted the manuscript. I.S., L.I.R., N.N., R.L.D., P.L.C., K.R.K. and I.W.D. provided critical revision of the manuscript for important intellectual content. All authors have approved the submitted version of the manuscript.

## References

1. Pagola-Lorz I, Vicente E, Ibáñez B, et al. Epidemiological study and genetic characterization of inherited muscle diseases in a northern Spanish region. Orphanet J Rare Dis. 2019;14(1):276.

2. Theadom A, Rodrigues M, Poke G, et al. A Nationwide, Population-Based Prevalence Study of Genetic Muscle Disorders. Neuroepidemiology. 2019;52(3-4):128–135.

3. Chulanova Y, Breier D, Peer D. Delivery of genetic medicines for muscular dystrophies. Cell Rep Med. 2025;6(1):101885.

4. Yeow D, Rudaks LI, Davis R, et al. Long-read sequencing for diagnosis of genetic myopathies. BMJ Neurol Open. 2025;7(1):e000990.

5. Erdmann H, Scharf F, Gehling S, et al. Methylation of the 4q35 D4Z4 repeat defines disease status in facioscapulohumeral muscular dystrophy. Brain. 2023;146(4):1388–1402.

6. Winckler PB, Chwal BC, Dos Santos MAR, et al. Diagnostic yield of multi-gene panel for muscular dystrophies and other hereditary myopathies. Neurol Sci. 2022;43(7):4473–4481.

7. Thuriot F, Gravel E, Buote C, et al. Molecular diagnosis of muscular diseases in outpatient clinics: A Canadian perspective. Neurol Genet. 2020;6(2):e408.

8. Beecroft SJ, Yau KS, Allcock RJN, et al. Targeted gene panel use in 2249 neuromuscular patients: the Australasian referral center experience. Ann Clin Transl Neurol. 2020;7(3):353–362.

9. Lee CL, Chuang CK, Chiu HC, et al. Application of whole exome sequencing in the diagnosis of muscular disorders: a study of Taiwanese pediatric patients. Front Genet. 2024;15:1365729.

10. Nicolau S, Milone M, Liewluck T. Guidelines for genetic testing of muscle and neuromuscular junction disorders. Muscle Nerve. 2021;64(3):255–269.

11. Kamsteeg EJ, Kress W, Catalli C, et al. Best practice guidelines and recommendations on the molecular diagnosis of myotonic dystrophy types 1 and 2. Eur J Hum Genet. 2012;20(12):1203–1208.

12. Giardina E, Camaño P, Burton-Jones S, et al. Best practice guidelines on genetic diagnostics of facioscapulohumeral muscular dystrophy: Update of the 2012 guidelines. Clin Genet. 2024;106(1):13–26.

13. Guruju NM, Jump V, Lemmers R, et al. Molecular Diagnosis of Facioscapulohumeral Muscular Dystrophy in Patients Clinically Suspected of FSHD Using Optical Genome Mapping. Neurol Genet. 2023;9(6):e200107.

14. Dai Y, Li P, Wang Z, et al. Single-molecule optical mapping enables quantitative measurement of D4Z4 repeats in facioscapulohumeral muscular dystrophy (FSHD). J Med Genet. 2020;57(2):109–120.

15. Zhang Q, Xu X, Ding L, et al. Clinical application of single-molecule optical mapping to a multigeneration FSHD1 pedigree. Mol Genet Genomic Med. 2019;7(3):e565.

16. Liu T, Conesa A. Profiling the epigenome using long-read sequencing. Nat Genet. 2025;57(1):27–41.

17. Stevanovski I, Chintalaphani SR, Gamaarachchi H, et al. Comprehensive genetic diagnosis of tandem repeat expansion disorders with programmable targeted nanopore sequencing. Sci Adv. 2022;8(9):eabm5386.

18. Ding Q, Balan J, Vidal-Folch N, et al. Rethinking the pathogenicity of intragenic DMD duplications detected by carrier screening: high prevalence of non-tandem duplications revealed by long-read sequencing. Genet Med. Published online 31 July 2025:101539.

19. Keegan NP, Wilton SD, Fletcher S. Breakpoint junction features of seven DMD deletion mutations. Hum Genome Var. 2019;6:39.

20. Startek M, Szafranski P, Gambin T, et al. Genome-wide analyses of LINE-LINE-mediated nonallelic homologous recombination. Nucleic Acids Res. 2015;43(4):2188–2198.

21. Lunt PW, Jardine PE, Koch MC, et al. Correlation between fragment size at D4F104S1 and age at onset or at wheelchair use, with a possible generational effect, accounts for much phenotypic variation in 4q35-facioscapulohumeral muscular dystrophy (FSHD). Hum Mol Genet. 1995;4(5):951–958.

22. Jones TI, Yan C, Sapp PC, et al. Identifying diagnostic DNA methylation profiles for facioscapulohumeral muscular dystrophy in blood and saliva using bisulfite sequencing. Clin Epigenetics. 2014;6(1):23.

23. Lemmers RJLF, van der Vliet PJ, Vreijling JP, et al. Cis D4Z4 repeat duplications associated with facioscapulohumeral muscular dystrophy type 2. Hum Mol Genet. 2018;27(20):3488–3497.

24. Lemmers RJLF, Butterfield R, van der Vliet PJ, et al. Autosomal dominant in cis D4Z4 repeat array duplication alleles in facioscapulohumeral dystrophy. Brain. 2024;147(2):414–426.

25. de Greef JC, Lemmers RJLF, van Engelen BGM, et al. Common epigenetic changes of D4Z4 in contraction-dependent and contraction-independent FSHD. Hum Mutat. 2009;30(10):1449–1459.

26. Lemmers RJLF, van der Vliet PJ, van der Gaag KJ, et al. Worldwide population analysis of the 4q and 10q subtelomeres identifies only four discrete interchromosomal sequence transfers in human evolution. Am J Hum Genet. 2010;86(3):364–377.

27. Fukuda H, Yamaguchi D, Nyquist K, et al. Father-to-offspring transmission of extremely long NOTCH2NLC repeat expansions with contractions: genetic and epigenetic profiling with long-read sequencing. Clin Epigenetics. 2021;13(1):204.

28. Yu J, Deng J, Guo X, et al. The GGC repeat expansion in NOTCH2NLC is associated with oculopharyngodistal myopathy type 3. Brain. 2021;144(6):1819–1832.

29. Ruggieri A, Naumenko S, Smith MA, et al. Multiomic elucidation of a coding 99-mer repeat-expansion skeletal muscle disease. Acta Neuropathol. 2020;140(2):231–235.

30. Deng J, Yu J, Li P, et al. Expansion of GGC Repeat in GIPC1 Is Associated with Oculopharyngodistal Myopathy. Am J Hum Genet. 2020;106(6):793–804.

31. Pan Y, Xue J, Chen J, et al. Assessment of GGC Repeat Expansion in GIPC1 in Patients with Parkinson’s Disease. Mov Disord. 2022;37(7):1557–1559.

32. Fan Y, Shen S, Yang J, et al. GIPC1 CGG Repeat Expansion Is Associated with Movement Disorders. Ann Neurol. 2022;91(5):704–715.

33. Eura N, Noguchi S, Ogawa M, et al. Complex associations of genetic/epigenetic variations of CGG repeats with patient phenotypes in oculopharyngodistal myopathy. medRxiv. Published online 15 May 2025:2025.05.13.25327490. doi:10.1101/2025.05.13.25327490

34. Yu J, Shan J, Yu M, et al. The CGG repeat expansion in RILPL1 is associated with oculopharyngodistal myopathy type 4. Am J Hum Genet. 2022;109(3):533–541.

35. Cortese A, Beecroft SJ, Facchini S, et al. A CCG expansion in ABCD3 causes oculopharyngodistal myopathy in individuals of European ancestry. Nat Commun. 2024;15(1):6327.

36. Jaganathan K, Kyriazopoulou Panagiotopoulou S, McRae JF, et al. Predicting Splicing from Primary Sequence with Deep Learning. Cell. 2019;176(3):535–548.e24.

37. Magri F, Brusa R, Bello L, et al. Limb girdle muscular dystrophy due to gene mutations: new mutations expand the clinical spectrum of a still challenging diagnosis. Acta Myol. 2020;39(2):67–82.

38. Huang M, Zhang Q, Jiao J, et al. Comprehensive genetic analysis of facioscapulohumeral muscular dystrophy by Nanopore long-read whole-genome sequencing. J Transl Med. 2024;22(1):451.

39. Yeetong P, Kulsirichawaroj P, Kumutpongpanich T, et al. Long-read Nanopore sequencing identified D4Z4 contractions in patients with facioscapulohumeral muscular dystrophy. Neuromuscul Disord. 2023;33(7):551–556.

40. Huang M, Zhang Q, Wu S, et al. Accurate detection of D4Z4 repeats, methylation and allele haplotype in facioscapulohumeral muscular dystrophy 1 using nanopore long-read adaptive sampling sequencing: a pilot study. J Med Genet. Published online 30 July 2025. doi:10.1136/jmg-2025-110827

41. Hiramuki Y, Kure Y, Saito Y, et al. Simultaneous measurement of the size and methylation of chromosome 4qA-D4Z4 repeats in facioscapulohumeral muscular dystrophy by long-read sequencing. J Transl Med. 2022;20(1):517.

42. Ishiura H, Shibata S, Yoshimura J, et al. Noncoding CGG repeat expansions in neuronal intranuclear inclusion disease, oculopharyngodistal myopathy and an overlapping disease. Nat Genet. 2019;51(8):1222–1232.

43. Ogasawara M, Iida A, Kumutpongpanich T, et al. CGG expansion in NOTCH2NLC is associated with oculopharyngodistal myopathy with neurological manifestations. Acta Neuropathol Commun. 2020;8(1):204.

44. Zhang T, Chancellor A, Liem B, et al. Neuronal intranuclear inclusion disease in New Zealand: A novel discovery. J Neurol Sci. 2024;460:122987.

45. Eura N, Iida A, Ogasawara M, Hayashi S, Noguchi S, Nishino I. RILPL1-related OPDM is absent in a Japanese cohort. Am J Hum Genet. 2022;109(11):2088–2089.

46. Scrimgeour EM, Mastaglia FL. Oculopharyngeal and distal myopathy: a case study from Papua New Guinea. Am J Med Genet. 1984;17(4):763–771.

47. Gushchin AG, Crum AV, Limbu BB, Quigley EP 3rd, Seward MS, Tabin GC. Simbu Ptosis: An Outreach Approach to Myogenic Ptosis in Eastern Highlands of Papua New Guinea-Experience and Results From a High-Volume Oculoplastic Surgical Camp. Ophthalmic Plast Reconstr Surg. 2017;33(2):139–143.

48. Payne A, Holmes N, Clarke T, Munro R, Debebe BJ, Loose M. Readfish enables targeted nanopore sequencing of gigabase-sized genomes. Nat Biotechnol. 2021;39(4):442–450.

49. Gamaarachchi H, Samarakoon H, Jenner SP, et al. Fast nanopore sequencing data analysis with SLOW5. Nat Biotechnol. 2022;40(7):1026–1029.

50. Samarakoon H, Ferguson JM, Gamaarachchi H, Deveson IW. Accelerated nanopore basecalling with SLOW5 data format. Bioinformatics. 2023;39(6). doi:10.1093/bioinformatics/btad352

51. Li H. Minimap2: pairwise alignment for nucleotide sequences. Bioinformatics. 2018;34(18):3094–3100.

52. Zheng Z, Li S, Su J, Leung AWS, Lam TW, Luo R. Symphonizing pileup and full-alignment for deep learning-based long-read variant calling. Nat Comput Sci. 2022;2(12):797–803.

53. Smolka M, Paulin LF, Grochowski CM, et al. Detection of mosaic and population-level structural variants with Sniffles2. Nat Biotechnol. 2024;42(10):1571–1580.

54. Jiang T, Liu Y, Jiang Y, et al. Long-read-based human genomic structural variation detection with cuteSV. Genome Biol. 2020;21(1):189.

55. Martin M, Patterson M, Garg S, et al. WhatsHap: fast and accurate read-based phasing. bioRxiv. Published online 2 November 2016. doi:10.1101/085050

56. Samarasinghe S, Deveson I, Gamaarachchi H. Base modification analysis in long read sequencing data using Minimod. bioRxiv. Published online 21 July 2025. doi:10.1101/2025.07.16.665072

57. McLaren W, Gil L, Hunt SE, et al. The Ensembl Variant Effect Predictor. Genome Biol. 2016;17(1):122.

58. Ziaei Jam H, Zook JM, Javadzadeh S, Park J, Sehgal A, Gymrek M. LongTR: genome-wide profiling of genetic variation at tandem repeats from long reads. Genome Biol. 2024;25(1):176.

59. Gasteiger E, Gattiker A, Hoogland C, Ivanyi I, Appel RD, Bairoch A. ExPASy: The proteomics server for in-depth protein knowledge and analysis. Nucleic Acids Res. 2003;31(13):3784–3788.

60. Weissensteiner H, Forer L, Kronenberg F, Schönherr S. mtDNA-Server 2: advancing mitochondrial DNA analysis through highly parallelized data processing and interactive analytics. Nucleic Acids Res. 2024;52(W1):W102–W107.

61. Cheng H, Concepcion GT, Feng X, Zhang H, Li H. Haplotype-resolved de novo assembly using phased assembly graphs with hifiasm. Nat Methods. 2021;18(2):170–175.

