## Supplementary Table 1 for "Targeted long-read sequencing enables comprehensive analysis of the genetic and epigenetic landscape of inherited myopathies"

**Supplementary Table 1. List of genetic loci for programmable targeted Oxford nanopore long-read sequencing myopathy panel.**

| Locus | Chromosome | Left Flank Position* | Right Flank Position* | Size (bp) |
|---|---|---|---|---|
| Mitochondrial DNA myopathies | | | | |
| Mitochondrial l | M | 1 | 16569 | 16569 |
| Facioscapulohumeral muscular dystrophy | | | | |
| 4q35 | 4 | 193360000 | 193572600 | 212601 |
| 10q26 | 10 | 134550000 | 134758134 | 208135 |
| *SMCHD1* | 18 | 2761308 | 3010649 | 249342 |
| *DNMT3B* | 20 | 34439113 | 34586082 | 146970 |
| *LRIF1* | 1 | 110911987 | 111028780 | 116794 |
| Short tandem repeat expansion myopathies | | | | |
| *DMPK* | 19 | 48547244 | 48660042 | 112799 |
| *CNBP* | 3 | 131862733 | 131978817 | 116085 |
| *PABPN1* | 14 | 17472305 | 17577200 | 104896 |
| *LRP12* | 8 | 105566729 | 105766466 | 199738 |
| *GIPC1* | 19 | 14554393 | 14672766 | 118374 |
| *NOTCH2NLC* | 1 | 148469507 | 148646912 | 177406 |
| *RILPL1* | 12 | 123418958 | 123582572 | 163615 |
| *NUTM2B-AS1/.* | 10 | 80472524 | 80799063 | 326540 |
| *ABCD3*[b] | 1 | 94216512 | 94416703 | 200192 |
| Dystrophinopathy (including putative modifier genes) | | | | |
| *DMD* | X | 30545824 | 33087553 | 2541730 |
| *SPP1* | 4 | 91252139 | 91359914 | 107776 |
| *LTBP4* | 19 | 43363474 | 43500376 | 136903 |
| *ANXA6* | 5 | 151587192 | 151744254 | 157063 |
| *CD40* | 20 | 47804319 | 47915915 | 111597 |
| *ACTN3* | 11 | 66492637 | 66609581 | 116945 |
| Emery-Dreiffus muscular dystrophy (including putative modifier genes) | | | | |
| *EMD* | X | 152565816 | 152668066 | 102251 |
| *LMNA* | 1 | 155171038 | 155328462 | 157425 |
| *SYNE1* | 6 | 153272981 | 153889113 | 616133 |
| *SYNE2* | 14 | 57919277 | 58484148 | 564872 |
| *FHL1* | X | 134405091 | 134569750 | 164660 |
| *TMEM43* | 3 | 14076960 | 14195625 | 118666 |
| *SUN1* | 7 | 869707 | 1050201 | 180495 |
| *SUN2* | 22 | 39154939 | 39314481 | 159543 |
| Limb-girdle muscular dystrophy | | | | |
| *DNAJB6* | 7 | 158473225 | 158655388 | 182164 |
| *TNPO3* | 7 | 130217065 | 130418043 | 200979 |
| *HNRNPDL* | 4 | 85702194 | 85810016 | 107823 |
| *CAPN3* | 15 | 40115533 | 40268365 | 152833 |
| *DYSF* | 2 | 71414648 | 71749464 | 334817 |
| *SGCA* | 17 | 50981282 | 51092985 | 111704 |
| *SGCB* | 4 | 55456644 | 55574238 | 117595 |
| *SGCG* | 13 | 22337744 | 22581974 | 244231 |
| *SGCD* | 5 | 156353192 | 157336763 | 983572 |
| *TCAP* | 17 | 40478914 | 40579814 | 100901 |
| *TRIM32* | 9 | 128830319 | 128944321 | 114003 |

| | | | | |
|---|---|---|---|---|
| *FKRP* | 19 | 49521623 | 49652810 | 131188 |
| *TTN* | 2 | 178958783 | 179363868 | 405086 |
| *POMT1* | 9 | 143665796 | 143786768 | 120973 |
| *ANO5* | 11 | 21870477 | 22453882 | 583406 |
| *FKTN* | 9 | 117682791 | 117878500 | 195710 |
| *POMT2* | 14 | 71434305 | 71580246 | 145942 |
| *POMGNT1* | 1 | 46015902 | 46147522 | 131621 |
| *DAG1* | 3 | 49447762 | 49614639 | 166878 |
| *PLEC* | 8 | 145023426 | 145185022 | 161597 |
| *TRAPPC11* | 4 | 186952463 | 187106791 | 154329 |
| *GMPPB* | 3 | 49696130 | 49803237 | 107108 |
| *CRPPA* | 7 | 16170757 | 16682215 | 511459 |
| *POGLUT1* | 3 | 122138674 | 122264425 | 125752 |
| *COL6A1* | 21 | 44306235 | 44430472 | 124238 |
| *COL6A2* | 21 | 44426193 | 44560938 | 134746 |
| *COL6A3* | 2 | 237765078 | 237955436 | 190359 |
| *LAMA2* | 6 | 130026331 | 130760229 | 733899 |
| *POMGNT2* | 3 | 43044562 | 43171424 | 126863 |
| *POPDC1* | 6 | 106222906 | 106362989 | 140084 |
| *POPDC3* | 6 | 106283653 | 106405749 | 122097 |
| *JAG2* | 14 | 99332841 | 99460749 | 127909 |
| Congenital muscular dystrophy | | | | |
| *COL12A1* | 6 | 76211502 | 76433203 | 221702 |
| *LARGE1* | 22 | 33577089 | 34430181 | 853093 |
| *RXYLT1* | 12 | 63708550 | 63838268 | 129719 |
| *B3GALNT2* | 1 | 234793084 | 234950322 | 157239 |
| *POMK* | 8 | 43312430 | 43450113 | 137684 |
| *B4GAT1* | 11 | 66289037 | 66391292 | 102256 |
| *DPM3* | 1 | 154229249 | 154329953 | 100705 |
| *DPM1* | 20 | 52655466 | 52779158 | 123693 |
| *DPM2* | 9 | 140092875 | 140195630 | 102756 |
| *DOLK* | 9 | 141098939 | 141201012 | 102074 |
| *ITGA7* | 12 | 55601197 | 55732679 | 131483 |
| *ITGA9* | 3 | 37403656 | 37877665 | 474010 |
| *CHKB* | 22 | 51039487 | 51161970 | 122484 |
| *GGPS1* | 1 | 234669063 | 234786246 | 117184 |
| *TRIP4* | 15 | 62147169 | 62314725 | 167557 |
| *LIMS2* | 2 | 128023635 | 128167027 | 143393 |
| *INPP5K* | 17 | 1333334 | 1455498 | 122165 |
| Metabolic myopathies & genetic rhabdomyolysis | | | | |
| *GYS1* | 19 | 51913010 | 52038209 | 125200 |
| *GAA* | 17 | 80951995 | 81070360 | 118366 |
| *AGL* | 1 | 99648594 | 99822284 | 173691 |
| *GBE1* | 3 | 81493135 | 81774407 | 281273 |
| *PYGM* | 11 | 64689618 | 64803279 | 113662 |
| *PFKM* | 12 | 48016476 | 48157749 | 141274 |
| *PHKA2* | X | 18425012 | 18616828 | 191817 |
| *PHKA1* | X | 70961736 | 71197893 | 236158 |

| | | | | |
|---|---|---|---|---|
| *PHKB* | 16 | 53207941 | 53548326 | 340386 |
| *PHKG2* | 16 | 31085292 | 31197812 | 112521 |
| *PGAM2* | 7 | 44171224 | 44274064 | 102841 |
| *LDHA* | 11 | 18442691 | 18556344 | 113654 |
| *ALDOA* | 16 | 30296810 | 30403102 | 106293 |
| *ENO3* | 17 | 4788440 | 4897445 | 109006 |
| *PGM1* | 1 | 63419205 | 63586042 | 166838 |
| *GYG1* | 3 | 151692127 | 151832489 | 140363 |
| *PGK1* | X | 76295795 | 76618161 | 322367 |
| *TPI1* | 12 | 6826416 | 6930246 | 103831 |
| *CPT2* | 1 | 53028234 | 53145633 | 117400 |
| *ACADVL* | 17 | 7068036 | 7176177 | 108142 |
| *ACADM* | 1 | 75511007 | 75674237 | 163231 |
| *ACADS* | 12 | 120664912 | 120779131 | 114220 |
| *HADHA* | 2 | 26175960 | 26330013 | 154054 |
| *HADHB* | 2 | 26228511 | 26375829 | 147319 |
| *SLC22A5* | 5 | 132839604 | 132965516 | 125913 |
| *ETFA* | 15 | 74036268 | 74232566 | 196299 |
| *ETFB* | 19 | 54383453 | 54505031 | 121579 |
| *ETFDH* | 4 | 161972873 | 162110372 | 137500 |
| *SLC25A32* | 8 | 104476285 | 104592838 | 116554 |
| *SLC25A20* | 3 | 48835896 | 48977909 | 142014 |
| *FLAD1* | 1 | 154072699 | 154182467 | 109769 |
| *PNPLA2* | 11 | 820350 | 927008 | 106659 |
| *ABHD5* | 3 | 43655696 | 43799958 | 144263 |
| *ALDH3A2* | 17 | 19546758 | 19684379 | 137622 |
| *LPIN1* | 2 | 11660753 | 11910525 | 249773 |
| *AMPD1* | 1 | 114634537 | 114757079 | 122543 |
| *MLIP* | 6 | 53719916 | 54156381 | 436466 |
| *OBSCN* | 1 | 227346855 | 227617714 | 270860 |
| *TANGO2* | 22 | 20344632 | 20494782 | 150151 |
| *SLC16A1* | 1 | 112874405 | 113019597 | 145193 |
| *TRAPPC2L* | 16 | 94876614 | 94981745 | 105132 |
| *PRKAG2* | 7 | 152678656 | 153099952 | 421297 |
| *RBCK1* | 20 | 400907 | 525908 | 125002 |
| | | Muscle channelopathies | | |
| *CACNA1S* | 1 | 200246459 | 200419435 | 172977 |
| *SCN4A* | 17 | 64759317 | 64893693 | 134377 |
| *KCNJ18* | 17 | 21591387 | 21703476 | 112090 |
| *KCNJ2* | 17 | 70996994 | 71108363 | 111370 |
| *KCNJ5* | 11 | 128876287 | 129006088 | 129802 |
| *CLCN1* | 7 | 144621527 | 144757513 | 135987 |
| *HSPG2* | 1 | 21596052 | 21811125 | 215074 |
| *LIFR* | 5 | 38673737 | 38907398 | 233662 |
| *KCNA1* | 12 | 4866952 | 4975267 | 108316 |
| | | Myofibrillar myopathies | | |
| *DES* | 2 | 219853171 | 219961530 | 108360 |
| *CRYAB* | 11 | 111868808 | 111983966 | 115159 |

| | | | | |
|---|---|---|---|---|
| *MYOT* | 5 | 138344284 | 138464287 | 120004 |
| *LDB3* | 10 | 87502666 | 87670190 | 167525 |
| *FLNC* | 7 | 130093304 | 130222199 | 128896 |
| *BAG3* | 10 | 120498963 | 120624379 | 125417 |
| *KY* | 3 | 137295261 | 137446989 | 151729 |
| *PYROXD1* | 12 | 21266314 | 21399929 | 133616 |
| *SVIL* | 10 | 29438432 | 29767597 | 329166 |
| *UNC45B* | 17 | 36045726 | 36187280 | 141555 |
| *MYL2* | 12 | 110840002 | 110950626 | 110625 |
| Nuclear gene mitochondrial myopathies | | | | |
| *ISCU* | 12 | 108481949 | 108588737 | 106789 |
| *MICU1* | 10 | 73188319 | 73547537 | 359219 |
| *MSTO1* | 1 | 154698770 | 154803498 | 104729 |
| *FDX2* | 19 | 10386158 | 10492125 | 105968 |
| *COX6A2* | 16 | 31765129 | 31865775 | 100647 |
| *SLC25A4* | 4 | 188436951 | 188544056 | 107106 |
| *TMEM126B* | 11 | 85515250 | 85623216 | 107967 |
| *OPA1* | 3 | 196238743 | 196444453 | 205711 |
| *POLG2* | 17 | 65297539 | 65416511 | 118973 |
| *PUS1* | 12 | 131927244 | 132043946 | 116703 |
| *YARS2* | 12 | 32553769 | 32682178 | 128410 |
| *LARS2* | 3 | 45354927 | 45621086 | 266160 |
| *AARS2* | 6 | 44082875 | 44197515 | 114641 |
| *ACAD9* | 3 | 131574316 | 131710789 | 136474 |
| *AGK* | 7 | 142817127 | 143021096 | 203970 |
| *AIFM1* | X | 128393471 | 128534685 | 141215 |
| *ATP5F1A* | 18 | 46221209 | 46345295 | 124087 |
| *C1QBP* | 17 | 5276513 | 5392569 | 116057 |
| *COX10* | 17 | 13927300 | 14189502 | 262203 |
| *COX6B1* | 19 | 38143302 | 38253762 | 110461 |
| *DGUOK* | 2 | 73885293 | 74017418 | 132126 |
| *DNA2* | 10 | 69233292 | 69391358 | 158067 |
| *FBXL4* | 6 | 99992904 | 100172374 | 179471 |
| *MGME1* | 20 | 17970141 | 18092263 | 122123 |
| *POLG* | 15 | 87021132 | 87139694 | 118563 |
| *POLRMT* | 19 | 521527 | 637843 | 116317 |
| *RRM2B* | 8 | 103280402 | 103414861 | 134460 |
| *MGME1* | 20 | 17970141 | 18092263 | 122123 |
| *SSBP1* | 7 | 143004160 | 143153741 | 149582 |
| *SLC25A42* | 19 | 19149813 | 19298846 | 149034 |
| *TK2* | 16 | 72252296 | 72396839 | 144544 |
| *TWNK* | 10 | 101820742 | 101927778 | 107037 |
| *TYMP* | 22 | 50986317 | 51090593 | 104277 |
| *TOP3A* | 17 | 18168234 | 18311813 | 143580 |
| *PNPLA8* | 7 | 109745069 | 109944274 | 199206 |
| *RNASEH1* | 2 | 3513959 | 3630861 | 116903 |
| *SDHA* | 5 | 159284 | 297721 | 138438 |
| *MPV17* | 2 | 27301915 | 27418103 | 116189 |

| | | | | |
|---|---|---|---|---|
| *COQ8A* | 1 | 226035448 | 226225910 | 190463 |
| *RMND1* | 6 | 152555351 | 152702800 | 147450 |
| *SCO2* | 22 | 50984133 | 51087026 | 102894 |
| *TAMM41* | 3 | 11736183 | 11892668 | 156486 |
| *SUCLA2* | 13 | 46916072 | 47308335 | 392264 |
| *SUCLG1* | 2 | 84375228 | 84511742 | 136515 |
| *TSFM* | 12 | 57701129 | 57826442 | 125314 |
| *NDUFB11* | X | 46501936 | 46604763 | 102828 |
| *GFER* | 16 | 1954138 | 2057694 | 103557 |
| *CHCHD10* | 22 | 24163028 | 24265165 | 102138 |
| *TMEM65* | 8 | 125388941 | 125555435 | 166495 |
| *MTO1* | 6 | 74587974 | 74736339 | 148366 |
| *MIEF2* | 17 | 18157403 | 18263010 | 105608 |
| *COQ2* | 4 | 86541628 | 86665010 | 123383 |
| *COQ6* | 14 | 68107579 | 68220719 | 113141 |
| Nemaline myopathies | | | | |
| *TPM3* | 1 | 153242532 | 153381881 | 139350 |
| *NEB* | 2 | 151887125 | 152236251 | 349127 |
| *ACTA1* | 1 | 228759946 | 228862804 | 102859 |
| *TPM2* | 9 | 35652644 | 35760710 | 108067 |
| *TNNT1* | 19 | 58176372 | 58293170 | 116799 |
| *KBTBD13* | 15 | 62836234 | 62939357 | 103124 |
| *CFL2* | 14 | 28858282 | 28963994 | 105713 |
| *KLHL40* | 3 | 42653548 | 42760555 | 107008 |
| *KLHL41* | 2 | 169936941 | 170053489 | 116549 |
| *LMOD3* | 3 | 69092877 | 69209846 | 116970 |
| *MYPN* | 10 | 68924932 | 69130820 | 205889 |
| *RYR3* | 15 | 31057434 | 31713766 | 656333 |
| Centronuclear myopathies | | | | |
| *MTM1* | X | 148786740 | 148990523 | 203784 |
| *DNM2* | 19 | 10794675 | 11010499 | 215825 |
| *BIN1* | 2 | 127432412 | 127591679 | 159268 |
| *CCDC78* | 16 | 676756 | 781128 | 104373 |
| *SPEG* | 2 | 219869623 | 220028369 | 158747 |
| *MAP3K20* | 2 | 173511969 | 173804582 | 292614 |
| *MTMR14* | 3 | 9591390 | 9744348 | 152959 |
| Core myopathies & other congenital myopathies | | | | |
| *RYR1* | 19 | 41185876 | 41441545 | 255670 |
| *SELENON* | 1 | 25587664 | 25705660 | 117997 |
| *MYH7* | 14 | 17563744 | 17686658 | 122915 |
| *MYH2* | 17 | 10378658 | 10507256 | 128599 |
| *MEGF10* | 5 | 127760566 | 128035524 | 274959 |
| *FXR1* | 3 | 183619912 | 183834535 | 214624 |
| *SECISBP2* | 9 | 101432274 | 101573440 | 141167 |
| *ACTN2* | 1 | 236015014 | 236225684 | 210671 |
| *CNTN1* | 12 | 40602132 | 41081590 | 479459 |
| *PAX7* | 1 | 18400908 | 18618930 | 218023 |
| *HACD1* | 10 | 17557536 | 17685876 | 128341 |

| | | | | |
|---|---|---|---|---|
| *STAC3* | 12 | 57161690 | 57269415 | 107726 |
| *MYL1* | 2 | 210720853 | 210845901 | 125049 |
| *TNNC2* | 20 | 47509705 | 47620239 | 110535 |
| *MYBPC1* | 12 | 101479451 | 101697245 | 217795 |
| *MYOD1* | 11 | 17767261 | 17869826 | 102566 |
| Multisystem proteinopathies | | | | |
| *VCP* | 9 | 35025243 | 35141804 | 116562 |
| *HNRNPA2B1* | 7 | 26258622 | 26386979 | 128358 |
| *HNRNPA1* | 12 | 54196770 | 54303665 | 106896 |
| *SQSTM1* | 5 | 180311594 | 180442429 | 130836 |
| *MATR3* | 5 | 139749827 | 139907737 | 157911 |
| *TIA1* | 2 | 70171187 | 70310402 | 139216 |
| *ANXA11* | 10 | 80970012 | 81124684 | 154673 |
| Congenital myasthenic syndromes | | | | |
| *CHRNA1* | 2 | 175186308 | 175326675 | 140368 |
| *CHRNB1* | 17 | 7299120 | 7411750 | 112631 |
| *CHRND* | 2 | 232962812 | 233073449 | 110638 |
| *CHRNE* | 17 | 4738020 | 4874783 | 136764 |
| *COLQ* | 3 | 15402540 | 15574153 | 171614 |
| *CHAT* | 10 | 50407901 | 50566753 | 158853 |
| *SYT2* | 1 | 201802402 | 202022645 | 220244 |
| *AGRN* | 1 | 400247 | 536183 | 135937 |
| *MUSK* | 9 | 122788435 | 123028207 | 239773 |
| *DOK7* | 4 | 3413090 | 3551231 | 138142 |
| *RAPSN* | 11 | 47547603 | 47658983 | 111381 |
| *GFPT1* | 2 | 69282145 | 69449608 | 167464 |
| *DPAGT1* | 11 | 119066886 | 119178712 | 111827 |
| *ALG2* | 9 | 111338129 | 111443646 | 105518 |
| *ALG14* | 1 | 94772745 | 94971349 | 198605 |
| *LRP4* | 11 | 46963618 | 47125510 | 161893 |
| *SNAP25* | 20 | 10211695 | 10400292 | 188598 |
| *COL13A1* | 10 | 70620064 | 70882777 | 262714 |
| *SLC5A7* | 2 | 108398100 | 108525570 | 127471 |
| *SLC18A3* | 10 | 50409116 | 50511526 | 102411 |
| *PREPL* | 2 | 44271600 | 44417164 | 145565 |
| *SLC25A1* | 22 | 19501495 | 19604689 | 103195 |
| *MYO9A* | 15 | 69591241 | 69985313 | 394073 |
| *VAMP1* | 12 | 6422621 | 6531325 | 108705 |
| *LAMB2* | 3 | 49099186 | 49211190 | 112005 |
| *TOR1AIP1* | 1 | 179187784 | 179330664 | 142881 |
| *UNC13A* | 19 | 17685571 | 17872500 | 186930 |
| *RPH3A* | 12 | 112497282 | 112925587 | 428306 |
| *LAMA5* | 20 | 64048904 | 64213029 | 164126 |
| *CHD8* | 14 | 15532671 | 15703516 | 170846 |
| *PURA* | 5 | 140582873 | 140700714 | 117842 |
| Other genetic myopathies | | | | |
| *FKBP14* | 7 | 30098289 | 30214384 | 116096 |
| *TNXB* | 6 | 31844357 | 32012566 | 168210 |

| | | | | |
|---|---|---|---|---|
| *PLOD1* | 1 | 11428324 | 11569657 | 141334 |
| *TAZ* | X | 152598030 | 152708213 | 110184 |
| *LAMP2* | X | 118751613 | 118894831 | 143219 |
| *VMA21* | X | 149614591 | 149727438 | 112848 |
| *STIM1* | 11 | 3869851 | 4208475 | 338625 |
| *ORAI1* | 12 | 121569745 | 121685891 | 116147 |
| *CASQ1* | 1 | 159277652 | 159388962 | 111311 |
| *ADSS1* | 14 | 98917108 | 99040211 | 123104 |
| *SMPX* | X | 21239285 | 21391420 | 152136 |
| *PLIN4* | 19 | 4435639 | 4552147 | 116509 |
| *GNE* | 9 | 36186447 | 36349058 | 162612 |
| *KLHL9* | 9 | 21293833 | 21399574 | 105742 |
| *ATP2A1* | 16 | 29108905 | 29234960 | 126056 |
| *CAV3* | 3 | 8674971 | 8883008 | 208038 |
| *CAVIN1* | 17 | 43209715 | 43330536 | 120822 |
| *HRAS* | 11 | 529238 | 634265 | 105028 |
| *MB* | 22 | 36015732 | 36147678 | 131947 |
| *SIL1* | 5 | 139422860 | 139869621 | 446762 |
| *TNNT3* | 11 | 1933743 | 2052750 | 119008 |
| *MYO18B* | 22 | 26153322 | 26543307 | 389986 |
| *DTNA* | 18 | 34635097 | 35133010 | 497914 |
| *MSTN* | 2 | 190495060 | 190602089 | 107030 |
| *DCST2* | 1 | 154107893 | 154223157 | 115265 |
| *DNAJB4* | 1 | 77767970 | 77906789 | 138820 |
| *TRIM63* | 1 | 25838705 | 25955842 | 117138 |
| *HMGCR* | 5 | 75767222 | 75892994 | 125773 |
| *HMGCS1* | 5 | 43490305 | 43616347 | 126043 |
| *P4HA1* | 10 | 73828593 | 74018486 | 189894 |
| *MYBPC3* | 11 | 47438226 | 47559533 | 121308 |
| *FAM111B* | 11 | 59006359 | 59126586 | 120228 |
| *GOSR2* | 17 | 47734521 | 47886955 | 152435 |
| *CHRNG* | 2 | 232976474 | 233084899 | 108426 |
| *LOXL4* | 10 | 99078069 | 99198321 | 120253 |
| *MYH3* | 17 | 10486098 | 10614858 | 128761 |
| *MYH8* | 17 | 10247824 | 10379456 | 131633 |
| *MYLPF* | 16 | 30707137 | 30814194 | 107058 |
| *COL4A1* | 13 | 109327772 | 109586621 | 258850 |
| *COL4A2* | 13 | 109485276 | 109793647 | 308372 |
| *VPS13A* | 9 | 89284011 | 89628104 | 344094 |
| *XK* | X | 37039348 | 37185703 | 146356 |
| *TRDN* | 6 | 124354293 | 124874889 | 520597 |
| *BET1* | 7 | 95148654 | 95290274 | 141621 |
| *DHX16* | 6 | 30467293 | 30587193 | 119901 |
| *TRPV1* | 17 | 3404480 | 3548442 | 143963 |
| *MYH14* | 19 | 53225697 | 53448071 | 222375 |
| *AHCY* | 20 | 35953380 | 36084908 | 131529 |
| *ASCC1* | 10 | 72916865 | 73138203 | 221339 |
| *ASCC3* | 6 | 101631937 | 102105319 | 473383 |

| | | | | |
|---|---|---|---|---|
| *ATP6V1A* | 3 | 116418238 | 116583252 | 165015 |
| *BICD2* | 9 | 104827924 | 104981391 | 153468 |
| *CHST14* | 15 | 38228334 | 38330478 | 102145 |
| *DYNC1H1* | 14 | 96150188 | 96342054 | 191867 |
| *EPG5* | 18 | 45988557 | 46208285 | 219729 |
| *HSPB1* | 7 | 77540142 | 77641764 | 101623 |
| *HSPB8* | 12 | 119111354 | 119262265 | 150912 |
| *TFG* | 3 | 103365050 | 103504713 | 139664 |
| *MAN2B1* | 19 | 12721018 | 12841240 | 120223 |
| *MPDU1* | 17 | 7437779 | 7547040 | 109262 |
| *MYMK* | 9 | 145678593 | 145792621 | 114029 |
| *PIEZO2* | 18 | 10778606 | 11361713 | 583108 |
| *PPP2R3C* | 14 | 29232399 | 29369427 | 137029 |
| *TTR* | 18 | 31697528 | 31839347 | 141820 |
| *GOLGA2* | 9 | 140412943 | 140533109 | 120167 |

*Left and right flank positions marked as 50 kbp on either side of each gene, except for *DMD*, for which flanking positions were marked as 150 kbp on either side of the gene. Coordinates based on T2T-chm13 (v2) reference genome.
