## Supplementary Files for "Targeted long-read sequencing enables comprehensive analysis of the genetic and epigenetic landscape of inherited myopathies"

**Supplementary Table 1. List of genetic loci for programmable targeted Oxford nanopore long-read sequencing myopathy panel.**

Attached as separate Excel file.

**Table S2**. **Comparison of uncommon *GIPC1* SNPs in newly diagnosed OPDM2 patients compared to a known Japanese OPDM2 haplotype**

| Japanese OPDM2 haplotype SNPs[10] (hg38 coordinates) | **L126** (European Australian) | | **L136** (Chinese) | | **L248** (New Zealand Māori) | | **L437** (New Zealand Māori, European Australian, Tongan) | |
|---|---|---|---|---|---|---|---|---|
| | STR expanded allele | Other allele | STR expanded allele | Other allele | STR expanded allele | Other allele | STR expanded allele | Other allele |
| chr19:14493342 T>A | A | A | A | T | A | A | A | A |
| chr19:14494246 T>C | C | C | C | C | C | C | C | C |
| chr19:14494369 G>A | A | G | A | G | A | G | A | G |
| chr19:14494383 C>T | T | C | T | C | T | T | T | T |
| chr19:14497761 G>A | G* | G | G* | G | A | G | A | G |
| chr19:14498125 G>A | A | A | A | G | A | A | A | A |
| chr19:14498399 A>G | G | G | G | A | G | G | G | G |

OPDM2; oculopharyngodistal myopathy type 2; SNPs, single nucleotide polymorphisms
*mismatches with the Japanese OPDM2 haplotype

**Demographics + previous testing on group 2 (Previously unsolved cases).**

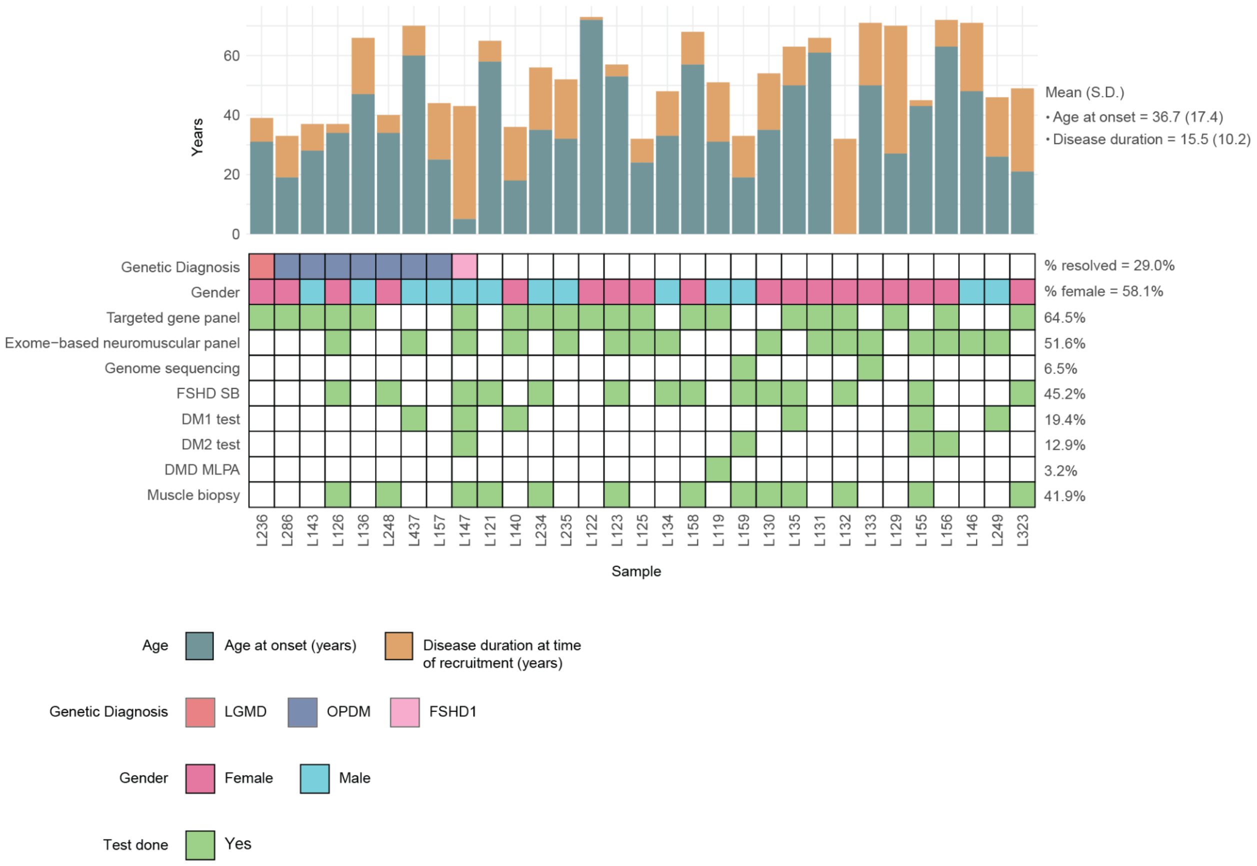


**Figure S1**. **Demographics and characteristics of participants with genetically unsolved or incompletely solved myopathies.** The upper bar chart shows the age of onset and the disease duration at time of recruitment. In the lower matrix, coloured boxes in the first row denote the diagnosis for each case following ONT LRS. Green boxes in the remaining rows indicate previous testing done in each participant.

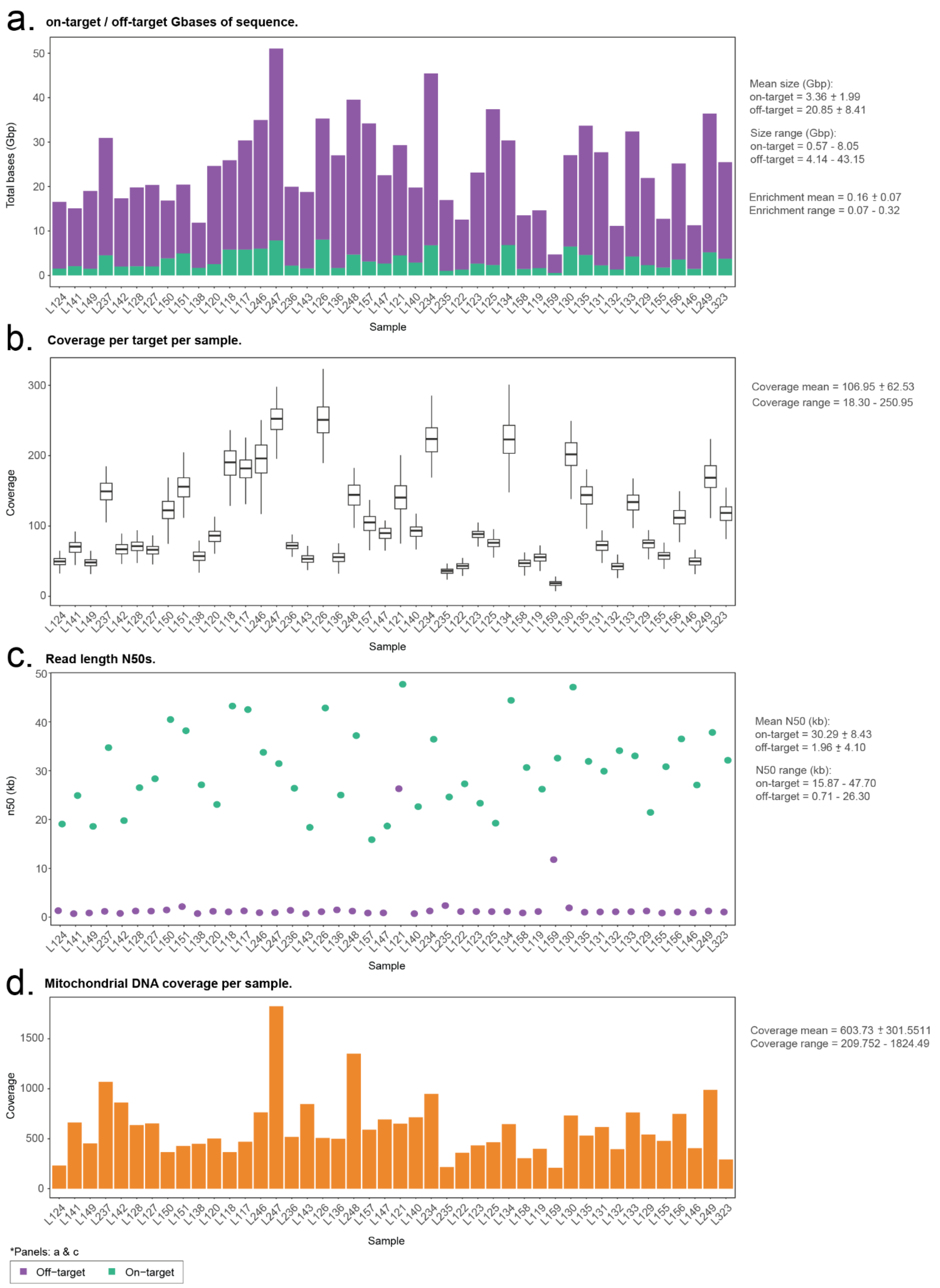


**Figure S2. Targeted ONT LRS myopathy panel sequencing metrics. (A)** Cohort sequencing data outputs showing the relative proportions of on-target (green) and off-target (purple) sequences in all individuals. **(B)** Distribution of coverage across the cohort. Each boxplot summarises the per-target coverage of an individual. **(C)** On-target (green) and off-target (purple) read length N50s distribution across the cohort. **(D)** Mitochondrial DNA coverage across all individuals.

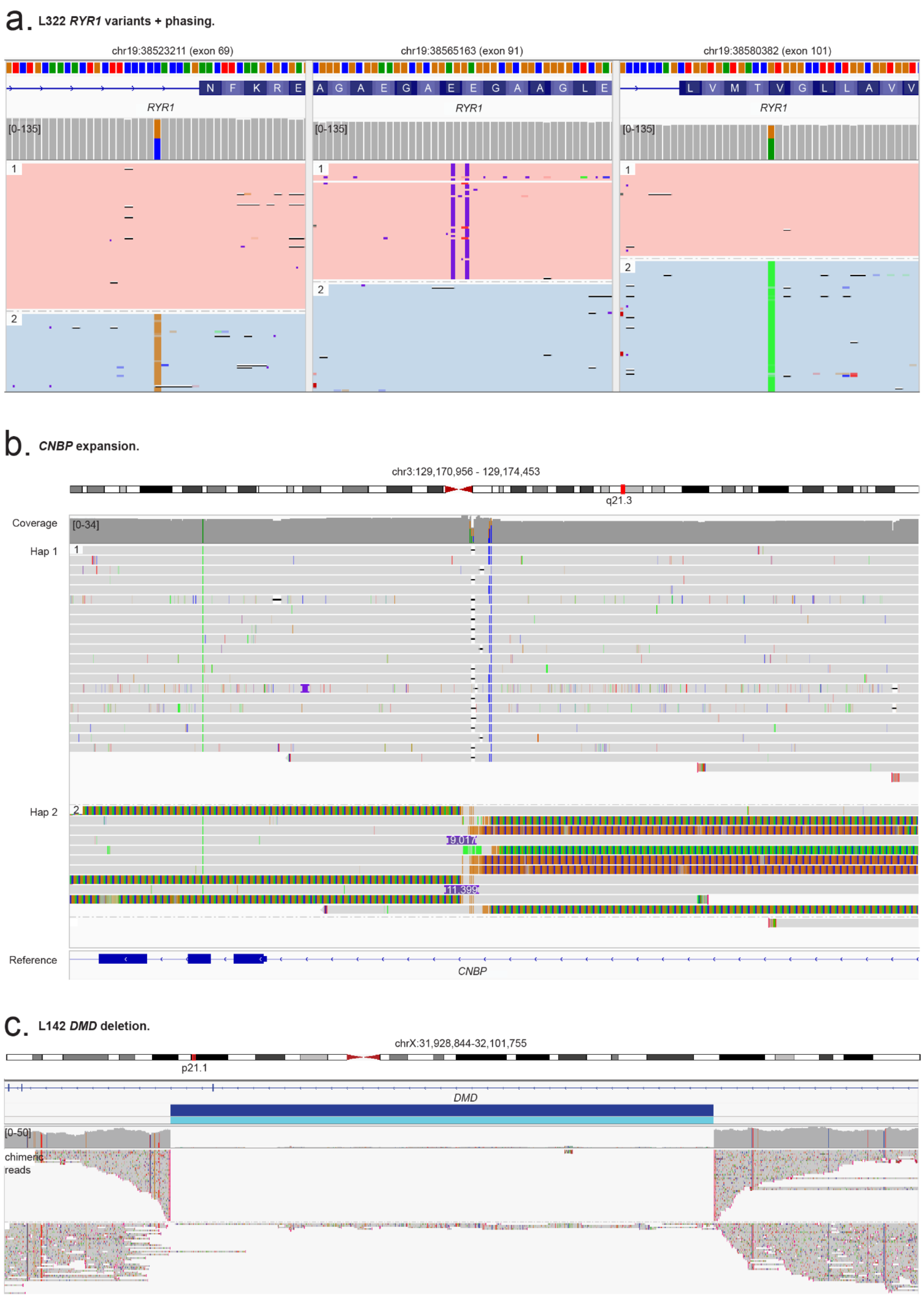


**Figure S3. Additional positive control genetic variants identified by ONT LRS. (A)** Case L141, with a diagnosis of congenital myopathy 1B, harbours three known variants in *RYR1*. ONT LRS identified that two variants (c.10348-6C>G and c.14524G>A) were on one allele while the third (c.12829G>T) was on the other allele without the need to sequence family members. **(B)** Case L237, with a diagnosis of myotonic dystrophy type 2, harbours a CCTG short tandem repeat expansion in *CNBP* with ONT LRS identifying mosaic in repeat length (2275–2870 repeats). **(C)** Case L142, with a known diagnosis of Duchenne muscular dystrophy, harbours a deletion of *DMD* exon 45. ONT LRS generated chimeric reads spanning the deletion and identified the exact breakpoints of the deletion: c.6439-94713_6614+8075del.

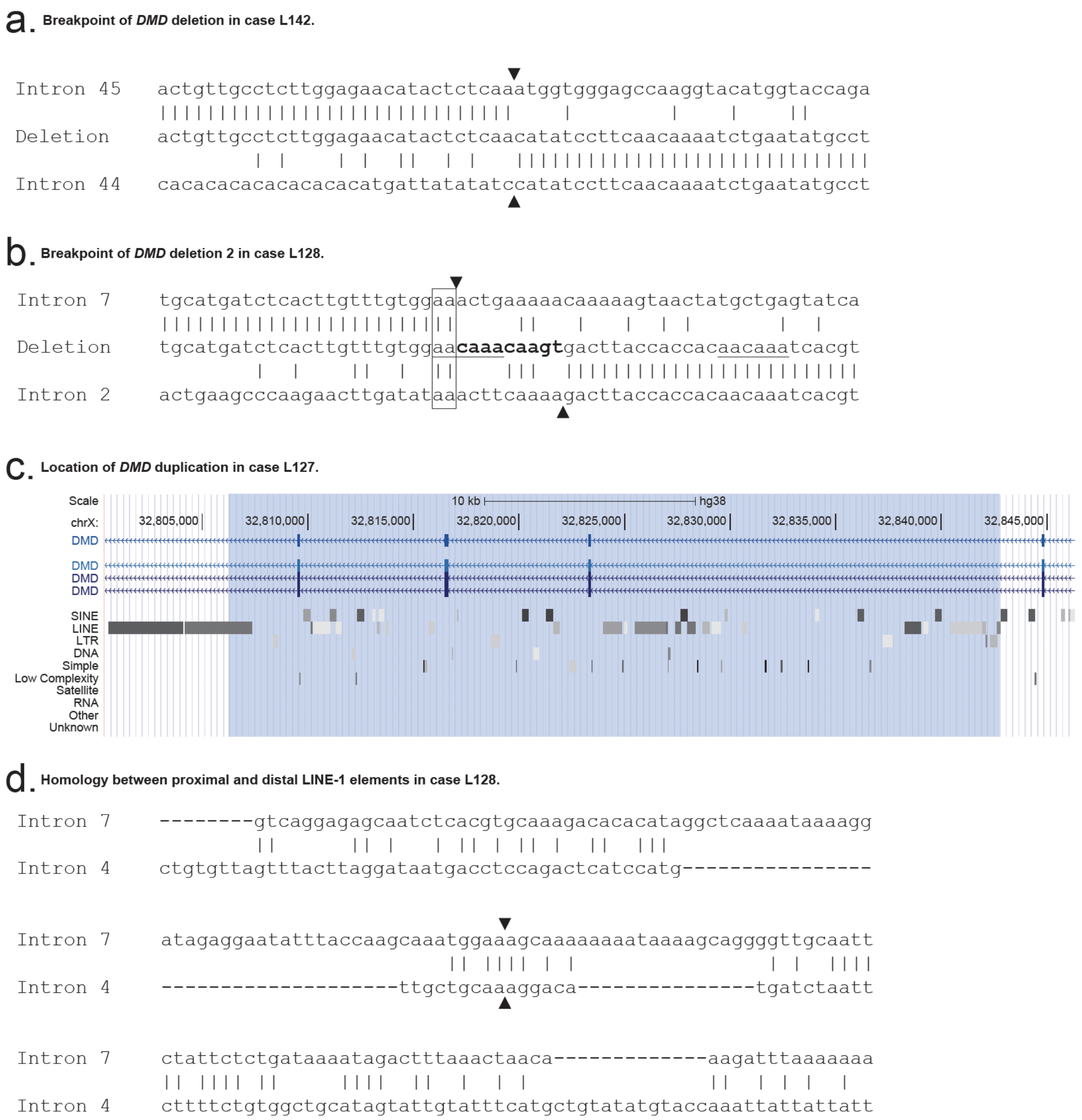


**Figure S4**. **ONT LRS characterises exact *DMD* copy number variation breakpoint junctions which informs potential mechanism of copy number variant generation. (A)** The deletion of exon 45 of *DMD* in case L142 was identified to have a clean breakend (arrowheads), consistent with non-homologous end-joining as a mechanism for the deletion.[11] **(B)** The deletion of exons 3–7 of *DMD* in case L128 was identified to have a novel 9 bp insertion (bold) at the deletion junction (arrowheads), part of which may be a copy of a nearby 4 bp tract (underlined). One of the deletion breakpoints is also adjacent to a 2 bp region of microhomology (boxed), suggestive of microhomology-mediated break induced replication as a possible mechanism.[11] **(C)** The two ends of the *DMD* exon 5–7 duplication in case L127 were found to be located within LINE-1 segments (highlighted in blue in the UCSC Genome Browser screenshot) indicating this duplication may have resulted from LINE-LINE-mediated nonallelic homologous recombination.[12] **(D)** The proximal and distal LINE-1 sequences in Case L127 demonstrate homology surrounding the breakpoint (arrowheads).

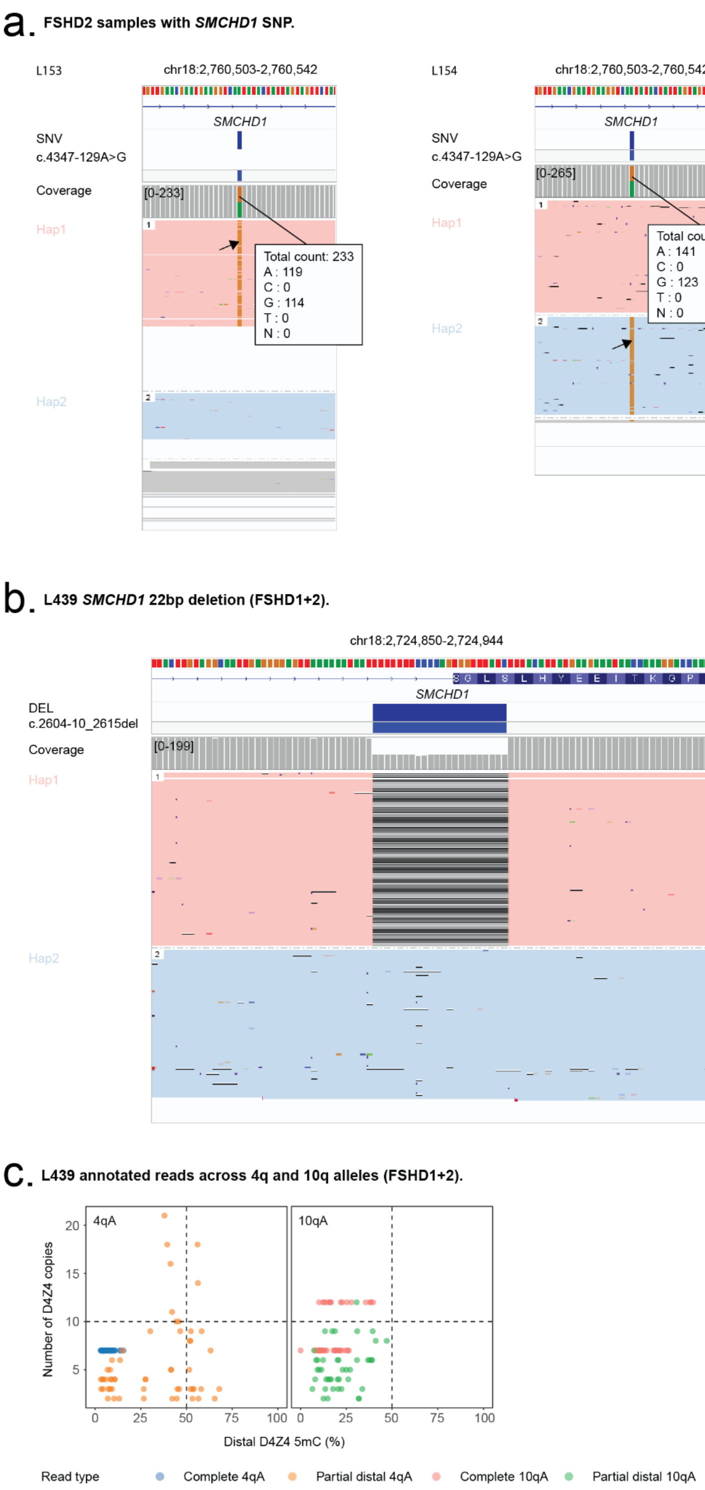


**Figure S5**. ***SMCHD1* variants in FSHD2 and FSHD1+2 cases and D4Z4 copy number and methylation analysis in FSHD1+2 case.** (**A**) Cases L438, L153 and L154, a mother and two daughters with known FSHD2, all harbour a deep intronic *SMCHD1* variant (c.4347-129A>G). (**B**) Case L439, a known case of FSHD1+2, ONT LRS identified the known likely pathogenic deletion in *SMCHD1* (c.2604-10_2615del). (**C**) Individual read 4qA and 10qA D4Z4 copy number and distal-most D4Z4 unit methylation plot in case L439 demonstrated severe hypomethylation of the contracted 7-copy D4Z4 4qA allele, milder hypomethylation of the non-contracted 4qA allele and hypomethylation of both 10qA alleles.

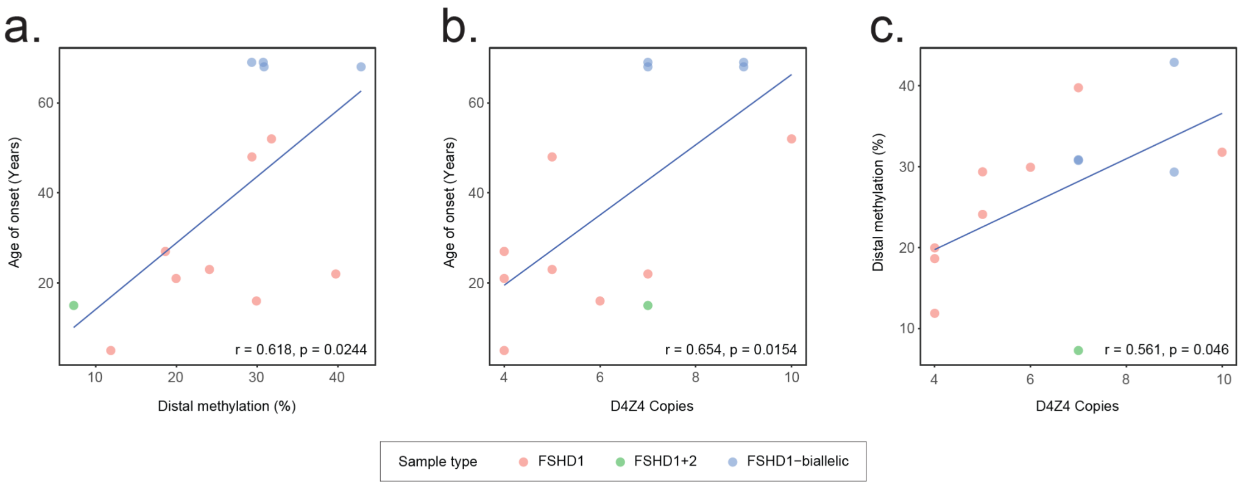


**Figure S6**. **Correlation between age of onset, D4Z4 copy number and D4Z4 methylation in FSHD1 and FSHD1+2 patients.** (**A**) Scatter plot of age of symptom onset and mean distal-most 4qA D4Z4 methylation showing positive correlation between distal-most D4Z4 unit methylation and age of onset. (**B**) Scatter plot of age of symptom onset and 4qA D4Z4 copy number showing positive correlation between 4qA D4Z4 copy number and age of onset. (**C**) Scatter plot of median distal-most 4qA D4Z4 unit methylation and 4qA D4Z4 copy number showing positive correlation between distal-most D4Z4 unit methylation and D4Z4 copy number. Pearson correlation coefficients were calculated to assess the linear association between variables. Corresponding p-values were derived using a two-sided test of no correlation.

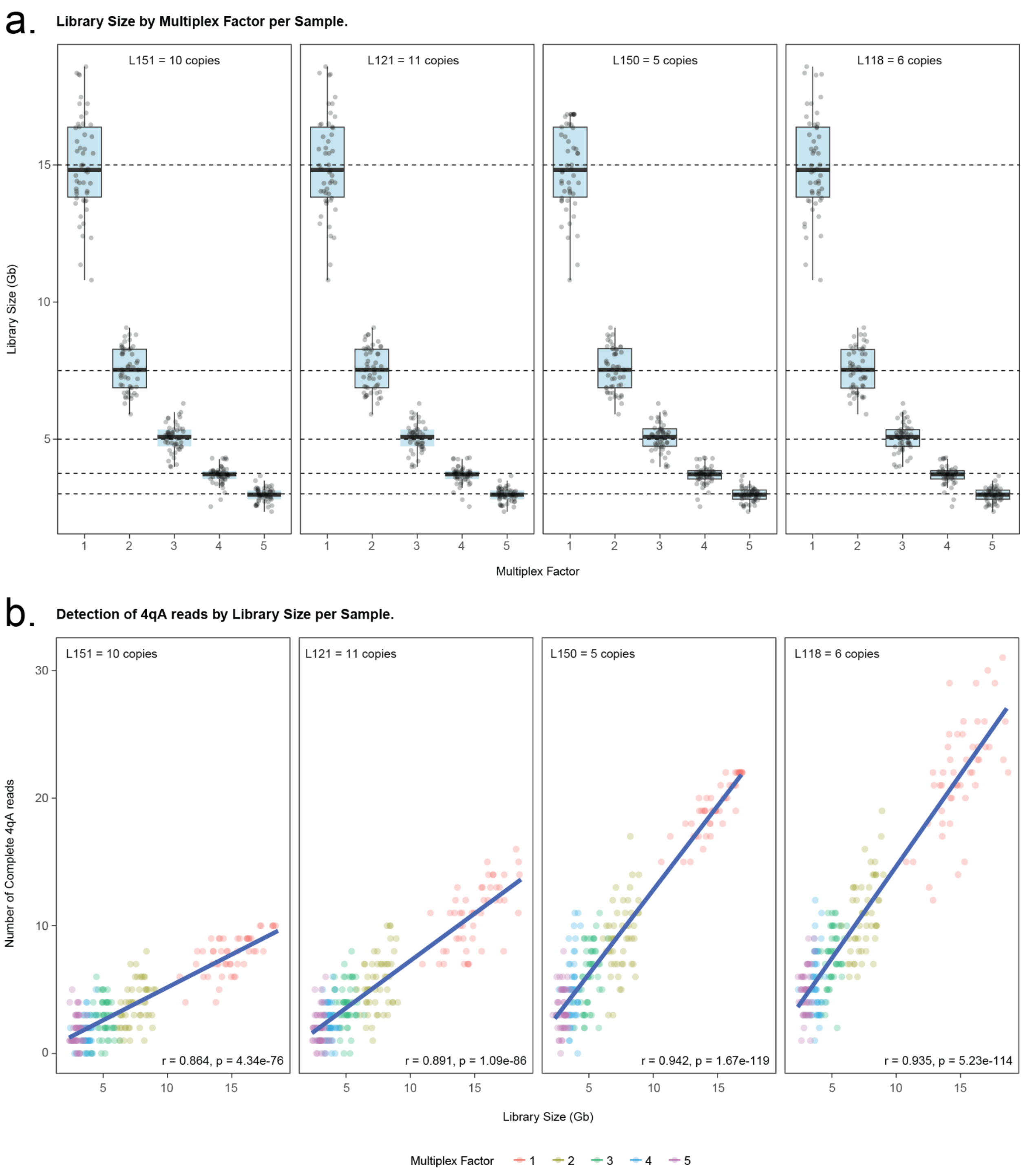


**Figure S7**. **FSHD downsampling experiments. (A)** Limit of detection analysis for library size. Each panel shows the library size distribution attained when simulating increasing multiplexing (sequencing the sample together with different numbers of samples) in representative individuals with varying 4qA D4Z4 copy numbers (5, 6, 10 & 11). **(B)** Limit of detection analysis for D4Z4 copy number. Each panel shows the number of complete 4qA reads when the library size increases. Trend lines indicate a positive correlation between increasing library sizes and number of complete 4qA reads found. Pearson correlation coefficients were calculated to assess the linear association between variables. Corresponding p-values were derived using a two-sided test of no correlation.

***NOTCH2NLC* Promoter methylation.**

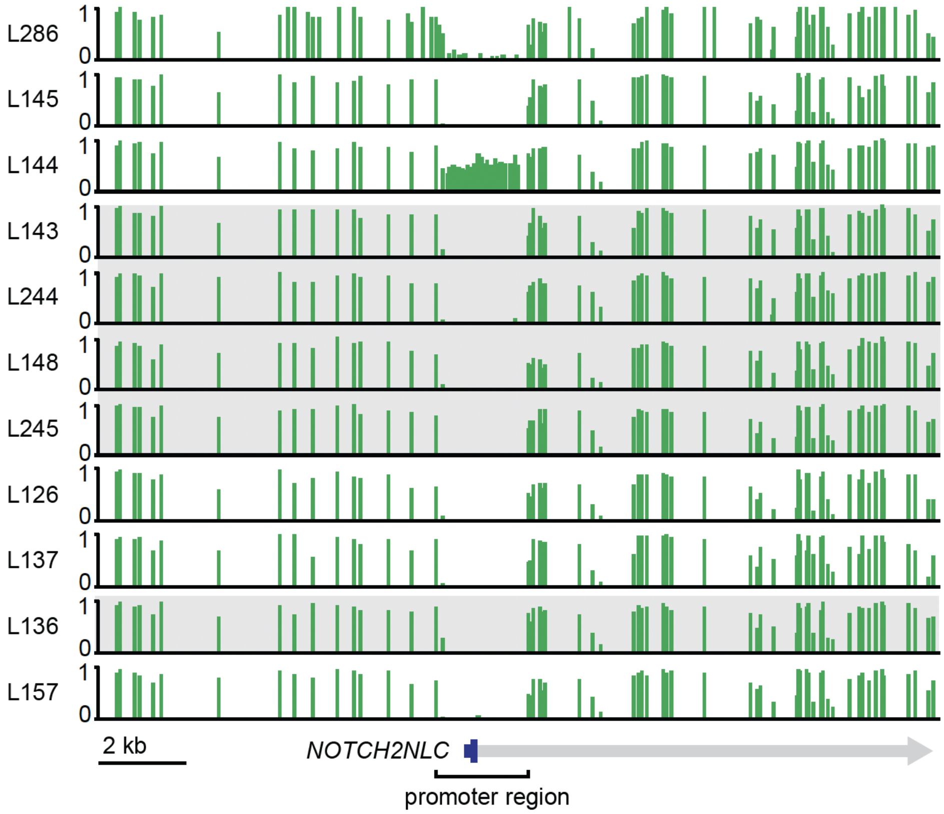


**Figure S8**. **CpG methylation across the *NOTCH2NLC* promoter in OPDM families.** Each row corresponds to an individual, with vertical bars showing methylation levels at different CpG sites. Case L144, who harbours a $(GGC)_{273–905}$ STR expansion in *NOTCH2NLC* but is asymptomatic, demonstrates hypermethylation of the *NOTCH2NLC* STR expansion site and promoter region. In comparison his daughter (L286), who harbours a $(GGC)_{130–621}$ STR expansion in *NOTCH2NLC* and has a diagnosis of OPDM2, as well as all other OPDM patients and their family members demonstrate little methylation in this region.

**a.** **L144 complex insertions in *PLIN4*.**

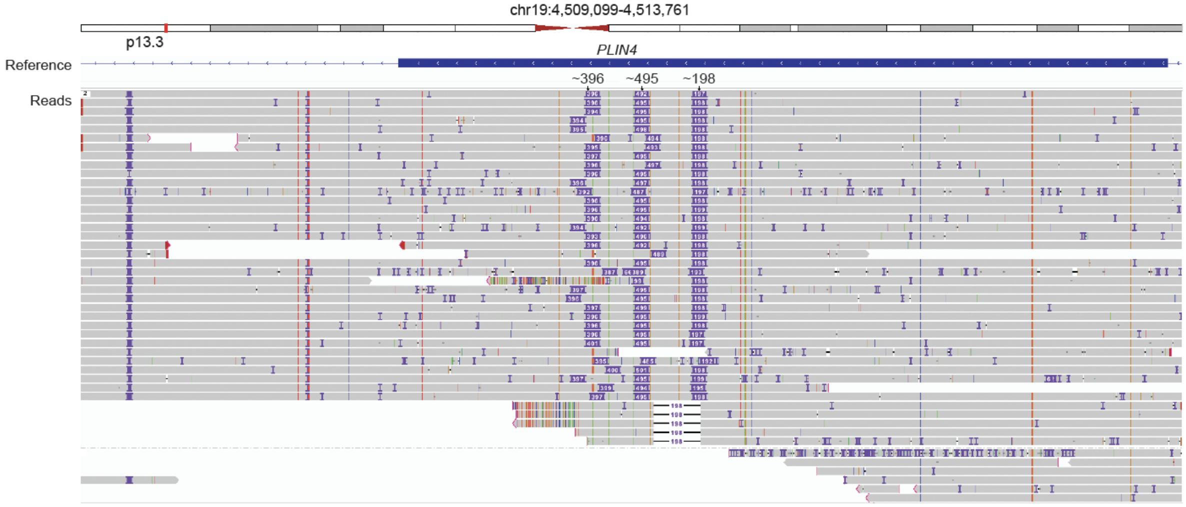


**b.** **PLIN4 amino acid sequence with L144/L286 insertions.**

MSAPDEGRRDPPKPKGKTLGSFFGSLPGFSSARNLVANAHSSARARPAADPTGAPAAEAAQPQAQVAAHPEQTAPWTEKELQP
SEKMVSGAKDLVCSKMSRAKDAVSS**GVASVVDVAKGVVQGGLDTTRSALTGTKEVVSSGVTGAMDMAKGAVQGGLDTSKAVLT**
**GTKDTVSTGLTGAVNVAKGTVQAGVDTTKTVLTGTKDTVTTGVMGAVNLAKGTVQTGVETSKAVLTGTKDAVSTGLTGAVNVA**
**RGSIQTGVDTSKTVLTGTKDTVCSGVTGAMNVAKGTIQTGVDTSKTVLTGTKDTVCSGVTGAMNVAKGTIQTGVDTSKTVLTG**
**TKDTVCSGVTGAMNVAKGTIQTGVDTTKTVLTGTKNTVCSGVTGAVNLAKEAIQGGLDTTKSMVMGTKDTMSTGLTGAANVAK**
**GAMQTGLNTTQNIATGTKDTVCSGVTGAMNLARGTIQTGVDTTKIVLTGTKDTVCSGVTGAANVAKGAVQGGLDTTKSVLTGT**
**KDAVSTGLTGAVNVAKGTVQTGVDTTKTVLTGTKDTVCSGVTSAVNVAKGAVQGGLDTTKSVVIGTKDTMSTGLTGAANVAKG**
**AVQTGVDTAKTVLTGTKDTVTTGLVGAVNVAKGTVQTGMDTTKTVLTGTKDTIYSGVTSAVNVAKGAVQTGLKTTQNIATGTK**
**NTFGSGVTSAVNVAKGAAQTGVDTAKTVLTGTKDTVTTGLMGAVNVAKGTVQTSVDTTKTVLTGTKDTVCSGVTGAANVAKGA**
**VQTGVDTAKTVLTGTKDTVCSGVTGAVNVAKGAVQTGLKTTQNIATGTKNTLGSGVTGAANVAK GAIQGGLDTTKSVLTGTKD**
**AVSTGLTGAVKLAKGTVQTGMDTTKTVLTGTKDAVCSGVTGAANVAKGAVQMGVDTAKTVLT GTKDTVTTGLMGAVNVAKGTV**
**QTSVDTTKTVLTGTKDTIYSGVTSAVNVAKGAVQTGLKTTQNIATGTKNTFGSGVTGAVNVAKGAVQTGVDTAKTVLTGTKDT**
**VTTGLMGAVNVAKGTVQTSVDTTKTVLTGTKDTVCSGVTGAANVAKGAVQTGVDTAKTVLT GTKDTVCSGVTGAANVAKGAVQ**
**TGLKTTQNIATGTKNTLGSGVTGAAKVAKGAVQGGLDTTKSVLTGTKD AVSTGLTGAVNLAKGTVQTGMDTTKTVLTGTKDAV**
**CSGVTGAANVAKGAVQTGVDTAKTVLTGTKDTVTTGLMGAVNVAKGTVQTSVDTTKTVLTGTKDTVCSGVTGAANVAKGAVQG**
**GLDTTKSVLTGTKDAVSTGLTGAVNLAKGTVQTGVDTSKTVLTGTKDTVCSGVTGAVNVAKGTVQTGVDTAKTVLSGAKDAVT**
**TGVTGAVNVAKGTVQTGVDASKAVLMGTKDTVFSGVTGAMSMAKGAVQGGLDTTKTVLTGTKDAVSA**GLMGSGNVATGATHTG
LSTFQNWLPSTPATSWGGLTSSRTTDNGGEQTALSPQEAPFSGISTPPDVLSVGPEPAWEAAATTKGLATDVATFTQGAAPGR
EDTGLLATTHGPEEAPRLAMLQNELEGLGDIFHPMNAEEQAQLAASQPGPKVLSAEQGSYFVRLGDLGPSFRQRAFEHAVSHL
QHGQFQARDTLAQLQDCFRLIEKAQQAPEGQPRLDQGSGASAEDAAVQEERDAGVLSRVCGLLRQLHTAYSGLVSSLQGLPAE
LQQPVGRARHSLCELYGIVASAGSVEELPAERLVQSREGVHQAWQGLEQLLEGLQHNPPLSWLVGPFALPAGGQ

**c.** **L144/286 PLIN4 repeat unit sequence alignment.**

1 GVASVVDVAKGVVQGGLDTTRSALTGTKEVVSS
2 GVTGAMDMAKGAVQGGLDTSKAVLTGTKDTVST
3 GLTGAVNVAKGTVQAGVDTTKTVLTGTKDTVTT
4 GVMGAVNLAKGTVQTGVETSKAVLTGTKDAVST
5 GLTGAVNVARGSIQTGVDTSKTVLTGTKDTVCS
6 GVTGAMNVAKGTIQTGVDTSKTVLTGTKDTVCS
7 GVTGAMNVAKGTIQTGVDTSKTVLTGTKDTVCS
8 GVTGAMNVAKGTIQTGVDTTKTVLTGTKNTVCS
9 GVTGAVNLAKEAIQGGLDTTKSMVMGTKDTMST
10 GLTGAANVAKGAMQTGLNTTQNIATGTKDTVCS
11 GVTGAMNLARGTIQTGVDTTKIVLTGTKDTVCS
12 GVTGAANVAKGAVQGGLDTTKSVLTGTKDAVST
13 GLTGAVNVAKGTVQTGVDTTKTVLTGTKDTVCS
14 GVTSAVNVAKGAVQGGLDTTKSVVIGTKDTMST
15 GLTGAANVAKGAVQTGVDTAKTVLTGTKDTVTT
16 GLVGAVNVAKGTVQTGMDTTKTVLTGTKDTIYS
17 GVTSAVNVAKGAVQTGLKTTQNIATGTKNTFGS
18 GVTSAVNVAKGAAQTGVDTAKTVLTGTKDTVTT
19 GLMGAVNVAKGTVQTSVDTTKTVLTGTKDTVCS
20 GVTGAANVAKGAVQTGVDTAKTVLTGTKDTVCS
21 GVTGAVNVAKGAVQTGLKTTQNIATGTKNTLGS
22 GVTGAANVAKGAIQGGLDTTKSVLTGTKDAVST
23 GLTGAVKLAKGTVQTGMDTTKTVLTGTKDAVCS
24 GVTGAANVAKGAVQMGVDTAKTVLTGTKDTVTT
25 GLMGAVNVAKGTVQTSVDTTKTVLTGTKDTIYS
26 GVTSAVNVAKGAVQTGLKTTQNIATGTKNTFGS
27 GVTGAVNVAKGAVQTGVDTAKTVLTGTKDTVTT
28 GLMGAVNVAKGTVQTSVDTTKTVLTGTKDTVCS
29 GVTGAANVAKGAVQTGVDTAKTVLTGTKDTVCS
30 GVTGAANVAKGAVQTGLKTTQNIATGTKNTLGS
31 GVTGAAKVAKGAVQGGLDTTKSVLTGTKDAVST
32 GLTGAVNLAKGTVQTGMDTTKTVLTGTKDAVCS
33 GVTGAANVAKGAVQTGVDTAKTVLTGTKDTVTT
34 GLMGAVNVAKGTVQTSVDTTKTVLTGTKDTVCS
35 GVTGAANVAKGAVQGGLDTTKSVLTGTKDAVST
36 GLTGAVNLAKGTVQTGVDTSKTVLTGTKDTVCS
37 GVTGAVNVAKGTVQTGVDTAKTVLSGAKDAVTT
38 GVTGAVNVAKGTVQTGVDASKAVLMGTKDTVFS
39 GVTGAMSMAKGAVQGGLDTTKTVLTGTKDAVSA

**Figure S9**. **Complex *n* × 99 bp repeat insertions in the *PLIN4* exon 5 VNTR in case L286 and L144. (A)** IGV browser view of the *PLIN4* exon 5 VNTR showing that, relative to the hg38 reference (which contains approximately 28 repeat units, about two fewer than most individuals), case L144 (and similarly L286, not shown) carries 11 additional 99 bp repeat units, visible as three large insertions (4 × 99 bp = 396 bp, 5 × 99 bp = 495 bp and 2 × 99 bp = 198 bp). **(B)** Amino acid sequence of the expanded PLIN4 allele generated by adding the three large insertions (396 bp, 495 bp and 198 bp) into their precise positions in exon 5 of the *PLIN4* MANE Select coding sequence and scanning for the correct open reading frame. Amino acids encoded by the full 39 × 99 bp VNTR are shown in bold, and those encoded by the inserted repeat units are highlighted in red. **(C)** Alignment of the 39 × 33 aa repeat units encoded by the expanded VNTR in cases L144 and L286. Red shading denotes amino acids derived from the inserted repeat units.

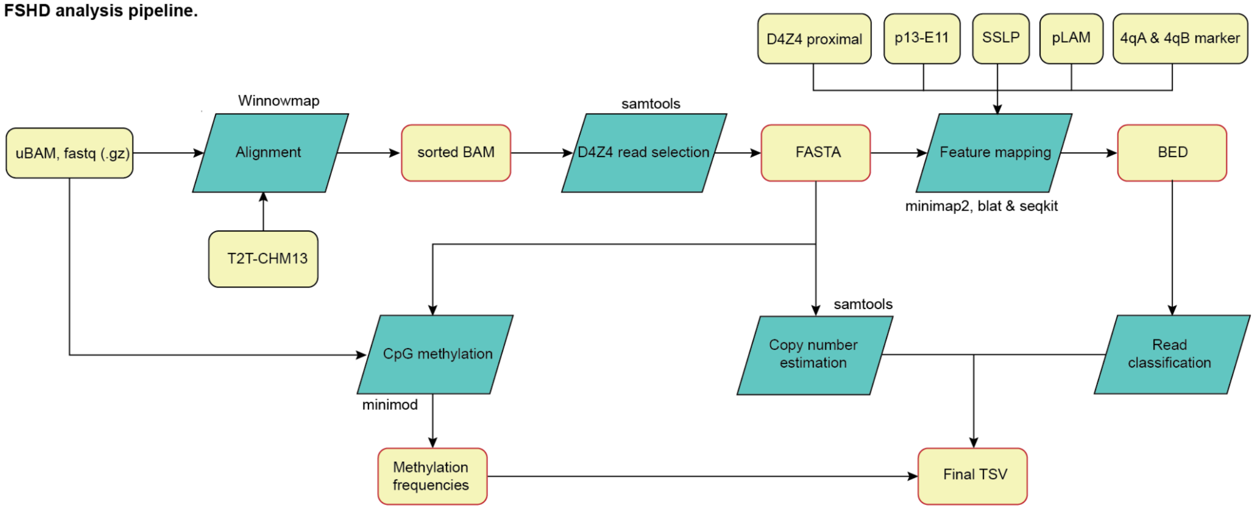


**Figure S10**. **Targeted ONT LRS FSHD analysis pipeline.** Flowchart of the newly developed FSHD analysis pipeline that uses LRS data to resolve the genetic and epigenetic architecture of FSHD. Inputs/outputs are represented by a yellow rounded rectangle, and processes represented by a green parallelogram. The process begins with aligning uBAM against the T2T-chm13 (v2) reference genome using Winnowmap, resulting in a sorted BAM file. The D4Z4 regions of chr4 and chr10 are then selected from the file and converted to a FASTA format. Each read is annotated with key features, including proximal elements (SSLP, p13-E11), the distal pLAM region, D4Z4 repeat units, and 4qA/4qB haplotype-specific marker using minimap2, blat and samtools, allowing the copy number estimation and classification of individual reads. Additionally, the FASTA file is mapped back to the uBAM for CpG methylation analysis with minimod, yielding methylation frequencies. Finally, all the genetic and epigenetic information are integrated into one final TSV output.

### Supplementary Methods

#### Skin Biopsy and Staining for INIs

For select participants and family members with OPDM-related STR expansions, a 3 mm punch biopsy of normal skin was performed. The specimens were fixed in 10% formalin solution, embedded in paraffin and cut into 4-mm thick sections. Sections were stained with hematoxylin and eosin. Immunohistochemistry was performed using antibodies against p62 (#610833, BD Sciences, 1:400) and ubiquitin (#Z0458, Dako, 1:1000).

#### *LAMA2* Sanger Sequencing and Reverse Transcription PCR (RT-PCR)

In case L236, who was found to carry a homozygous intronic VUS in *LAMA2*, segregation studies were performed in the proband and her mother using primers (5′-CTTGTCTGGCTCCTGTCTGA-3′; 5′-ATTTGGAAAACAGTGATGCTTCA-3′) that amplify *LAMA2* exon 19 and intron 19 (chr6:129287952–129288286 [hg38]) to produce a 335 bp amplicon. Amplicons underwent Sanger sequencing using Applied Biosystems BigDye Terminator cycle sequencing protocols. Sequences were visualised and analysed using Snapgene Software V7.1.

Skin biopsy-derived primary fibroblast lines were established from case L236 and an unrelated control individual. Fibroblast cell lines were maintained in culture medium comprising Dulbecco's Modified Eagle Medium (DMEM), 10% (v/v) fetal bovine serum (FBS), 1% (v/v) penicillin-streptomycin and 1% (v/v) L-glutamine (Gibco, Life Technologies). Fibroblasts were maintained in 5% $CO_2$ humidified air at 37°C. RNA extraction from fibroblasts was conducted using RNEasy Mini Kit (Qiagen) and reverse-transcribed template was prepared using iScript cDNA Synthesis Kit (Bio-Rad). PCR amplification of cDNA was conducted using a forward primer designed to anneal to the exon-exon boundary between *LAMA2* exons 17 and 18 (5′-CCCATCCAATAACTTTAGCCCA-3′), and a reverse primer designed to anneal to exon 21 (5′-GCGGTCACATTTCTTCCCTG-3′). The RT-PCR amplicons were size fractionated using a 2% (w/v) agarose gel, gel-purified using Isolate II PCR and Gel Kit (Bioline) and then Sanger sequenced using Applied Biosystems BigDye Terminator cycle sequencing protocols. Sequences were visualised and analysed using Snapgene Software V7.1.

**Supplementary References**


1. Stender S, Chakrabarti RS, Xing C, Gotway G, Cohen JC, Hobbs HH. Adult-onset liver disease and hepatocellular carcinoma in S-adenosylhomocysteine hydrolase deficiency. Mol Genet Metab. 2015;116(4):269-274.
2. Baric I, Fumic K, Glenn B, et al. S-adenosylhomocysteine hydrolase deficiency in a human: a genetic disorder of methionine metabolism. Proc Natl Acad Sci U S A. 2004;101(12):4234-4239.
3. Ghaoui R, Cooper ST, Lek M, et al. Use of whole-exome sequencing for diagnosis of limb-girdle muscular dystrophy: outcomes and lessons learned. JAMA Neurol. 2015;72(12):1424-1432.
4. Sullivan PJ, Quinn JMW, Ajuyah P, Pinese M, Davis RL, Cowley MJ. Data-driven insights to inform splice-altering variant assessment. Am J Hum Genet. 2025;112(4):764-778.
5. Ronchi D, Garone C, Bordoni A, et al. Next-generation sequencing reveals DGUOK mutations in adult patients with mitochondrial DNA multiple deletions. Brain. 2012;135(Pt 11):3404-3415.
6. Ruggieri A, Naumenko S, Smith MA, et al. Multiomic elucidation of a coding 99-mer repeat-expansion skeletal muscle disease. Acta Neuropathol. 2020;140(2):231-235.
7. Yang K, Zeng YH, Qiu YS, et al. Expanding the phenotype and genotype spectra of PLIN4-associated myopathy with rimmed ubiquitin-positive autophagic vacuolation. Acta Neuropathol. 2022;143(6):733-735.
8. Llansó L, Stevanovski I, Morís G, et al. Repeat expansions in PLIN4 cause autosomal dominant vacuolar myopathy with sarcolemmal features. Ann Clin Transl Neurol. 2025;12(10):2136-2151.
9. Carnazzi A, Iannibelli E, Gibertini S, et al. Genetic variability in the PLIN4 gene: A new sequence duplication causing autophagic vacuolar myopathy. Genes Dis. Published online 10 Sep 2025:101849. doi.org/10.1016/j.gendis.2025.101849.
10. Eura N, Noguchi S, Ogawa M, et al. Complex associations of genetic/epigenetic variations of CGG repeats with patient phenotypes in oculopharyngodistal myopathy. medRxiv. Published online 15 May 2025:2025.05.13.25327490. doi:10.1101/2025.05.13.25327490
11. Keegan NP, Wilton SD, Fletcher S. Breakpoint junction features of seven DMD deletion mutations. Hum Genome Var. 2019;6:39.
12. Startek M, Szafranski P, Gambin T, et al. Genome-wide analyses of LINE-LINE-mediated nonallelic homologous recombination. Nucleic Acids Res. 2015;43(4):2188-2198
